# Understanding the effectiveness of government interventions in Europe’s second wave of COVID-19

**DOI:** 10.1101/2021.03.25.21254330

**Authors:** Mrinank Sharma, Sören Mindermann, Charlie Rogers-Smith, Gavin Leech, Benedict Snodin, Janvi Ahuja, Jonas B. Sandbrink, Joshua Teperowski Monrad, George Altman, Gurpreet Dhaliwal, Lukas Finnveden, Alexander John Norman, Sebastian B. Oehm, Julia Fabienne Sandkühler, Thomas Mellan, Jan Kulveit, Leonid Chindelevitch, Seth Flaxman, Yarin Gal, Swapnil Mishra, Jan Markus Brauner, Samir Bhatt

**Author notes:** Equal contribution to first authorship (Mrinank Sharma, Sören Mindermann, Jan Markus Brauner). Equal contribution to senior authorship (Swapnil Mishra, Jan Markus Brauner, Samir Bhatt).

## Abstract

As European governments face resurging waves of COVID-19, non-pharmaceutical interventions (NPIs) continue to be the primary tool for infection control. However, updated estimates of their relative effectiveness have been absent for Europe’s second wave, largely due to a lack of collated data that considers the increased subnational variation and diversity of NPIs. We collect the largest dataset of NPI implementation dates in Europe, spanning 114 subnational areas in 7 countries, with a systematic categorisation of interventions tailored to the second wave. Using a hierarchical Bayesian transmission model, we estimate the effectiveness of 17 NPIs from local case and death data. We manually validate the data, address limitations in modelling from previous studies, and extensively test the robustness of our estimates. The combined effect of all NPIs was smaller relative to estimates from the first half of 2020, indicating the strong influence of safety measures and individual protective behaviours--such as distancing--that persisted after the first wave. Closing specific businesses was highly effective. Gathering restrictions were highly effective but only for the strictest limits. We find smaller effects for closing educational institutions compared to the first wave, suggesting that safer operation of schools was possible with a set of stringent safety measures including testing and tracing, preventing mixing, and smaller classes. These results underscore that effectiveness estimates from the early stage of an epidemic are measured relative to pre-pandemic behaviour. Updated estimates are required to inform policy in an ongoing pandemic.

## Introduction

The first wave of the novel coronavirus, SARS-CoV-2, resulted in marked excess mortality across most Western European countries. Most of these countries implemented a suite of unprecedented non-pharmaceutical interventions (NPIs) including the closure of businesses, schools, and bans on gatherings (*1*–*1*). Although partial control was achieved in the summer months, a second wave (*5, 6*) of the epidemic followed the reopening of European societies around September 2020. With vaccines set to reach only a minority of the global population in 2021 (*1*) and current vaccine roll-out delays in Europe, NPIs remain the primary tool for infection control in the short term. Furthermore, new variants of concern with mutations that contribute to higher transmissibility, severity, or antigenic escape (*1*–*1*) further increase the need for targeted methods to control infections while reducing the social and economic consequences of broad lockdowns.

The relative effectiveness of NPIs in the first wave of the pandemic has been determined by relating the implemented NPIs to the course of the pandemic in different countries (*1*–*3, 11*–*1*). However, NPI effectiveness estimates from the second wave are expected to be substantially more representative of future waves and thus more relevant to government decision making, for several reasons. The statistical models used to estimate NPI effectiveness rely on comparing epidemiological observations in the presence and absence of NPIs. However, disease transmission in the absence of a given NPI depends strongly on behaviour. Contact patterns changed with the arrival of the first wave, as individuals adopted voluntary practices to reduce infection including physical distancing, avoiding crowded places, mask-wearing, and working from home (*1*), while organizations adopted ongoing safety measures such as testing and separating cohorts in schools (*1*). Recent polling suggests that behavioural patterns have comparatively stabilized since the summer of 2020 (*1*), indicating that as long as these behavioural changes stay in place, effect sizes in the second wave are more relevant to potential further waves.

Furthermore, many NPIs in the first wave started within a short period in March 2020, which made it challenging to disentangle the effects of individual NPIs (*1, 2*). European governments have also adopted a more granular selection of interventions in the second wave, such as time-staggered closure of different business types. This stratification was absent in the first wave. Moreover, NPIs were implemented subnationally, which greatly increases the number of data points available for this study. Finally, in the first wave, the data quality was affected by pervasive undercounting and sampling bias in the case data, and the variable time lag between infection and death (*1*) reduced the statistical precision of mortality data (*1*). Taken together, the data from the second wave is predicted to give a much more detailed, robust, and contemporary picture of the impact of specific interventions.

Here, we provide the first effect estimates for individual interventions during Europe’s second wave. Countries typically implemented several NPIs concurrently, or in grouped tiers (*1*–*1*), making their individual effects non-identifiable. A multinational dataset is required to overcome this limitation since different countries implemented different sets of NPIs at different times. We also require subnational data since modelling on a national scale would obscure local heterogeneity from transmission timings (*21, 22*), socioeconomic heterogeneity, and differing policy decisions. Ignoring this subnational variation can result in ecological fallacies (*1*) and biased effect estimates. Infection heterogeneity preceding the second wave in the UK is a salient example, as there was a strong north/south geographic divide meaning that analysis on a national scale would have obscured increases in transmission due to aggregation. Modelling efforts for the second wave thus require a novel NPI dataset, since the utility of existing trackers is limited by a lack of granular subnational data, inconsistencies (*1*), missing entries (*1*), and broad intervention definitions ill-suited for the second wave.

We introduce a bespoke and systematic categorisation of interventions and estimate effect sizes across a randomized sample of 114 regions in 7 European countries (Austria, the Czech Republic, England, Germany, Italy, the Netherlands, Switzerland). We manually collected the intervention data with several validation procedures to ensure high quality and consistency. Using a semi-mechanistic Bayesian transmission model with a latent stochastic process, we estimate effects for all NPIs jointly from case and death data. Since estimates of NPI effectiveness can be highly sensitive to modelling decisions (*14, 26*), we evaluate robustness to changes in the data, model, epidemiological assumptions, and potential unobserved confounding factors.

### Differences between Europe’s first and second wave

A key challenge for distinguishing the effects of specific interventions is that governments often implement several NPIs simultaneously (Fig. 1). During the first wave, interventions were implemented within a short time window: for a given intervention and region, on average 83% of the other interventions in that region started in the same 10 day period (*1*). In the second wave, NPI implementation was spaced out (Fig. 1), with only 23% of interventions starting in the same 10 day period. The increased temporal spacing combined with an even larger and subnational dataset (9.2x more NPI implementations than the largest study that focused on Europe (*1*)) allow us to disentangle the effects of 17 individual NPIs. For most NPIs, we were able to robustly isolate their individual effects because their collinearity was limited and our dataset large. Specifically, for each pair of NPIs that we are able to disentangle, we observed one without the other for 6969 region-days on average (with a minimum of 635 region-days for limiting household mixing in private to ≤10 attendees and to ≤30 attendees). However, we only show the combined effect of indoor and outdoor gathering bans (of various stringencies) since these comprise all 6 NPI pairs that score lowest on the aforementioned metric. Our estimates are robust to changes in data and model parameters (see *Robustness of estimates*), in contrast to smaller datasets on first-wave data (*1, 26*), indicating (*1*) that the data are sufficient to overcome collinearity.

**Fig. 1.**
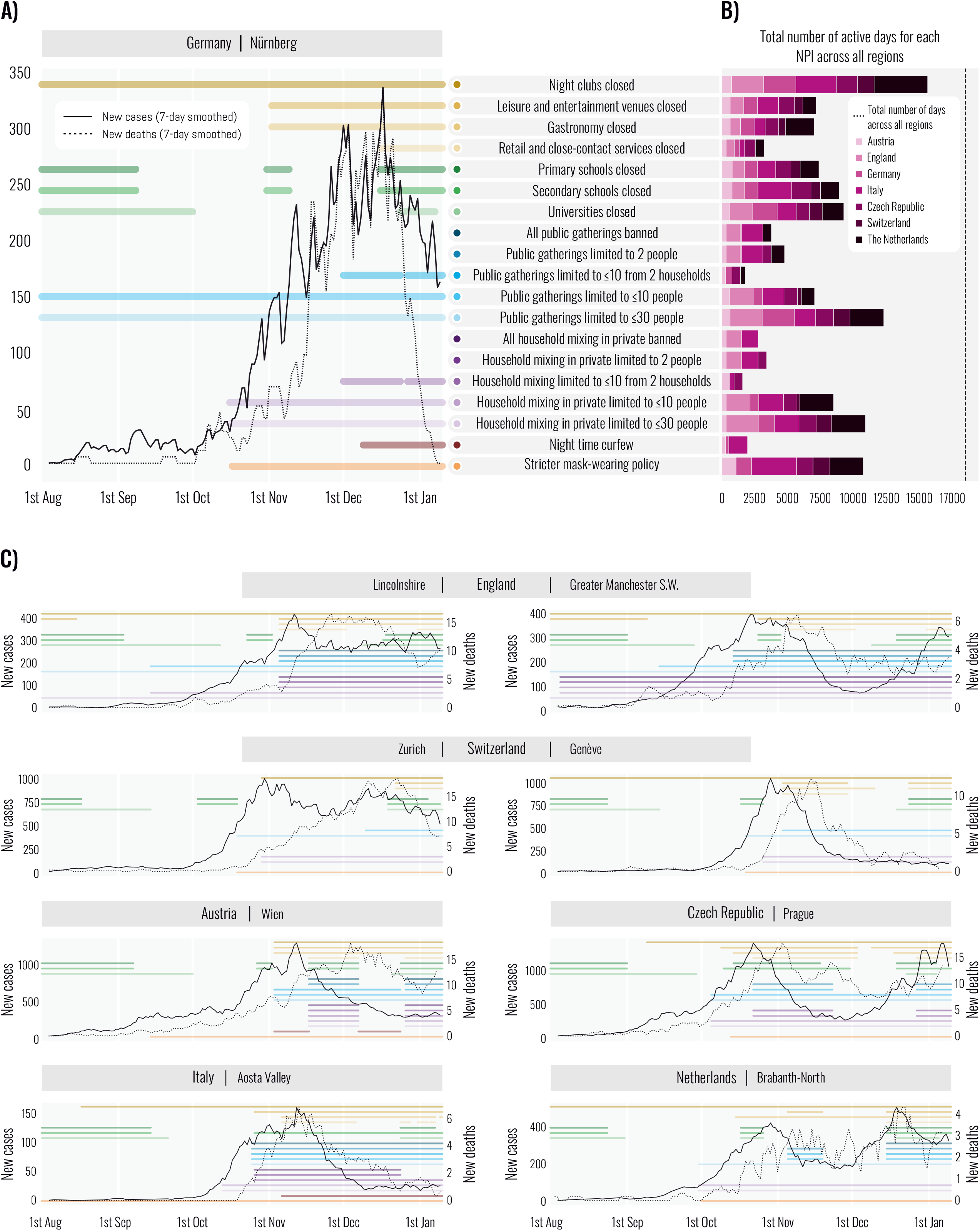
Dataset. **(A)** Cases, deaths and implementation dates of nonpharmaceutical interventions in an example region (Nürnberg, Germany). Colored lines indicate the dates that each intervention was active. Colors represent different interventions. **(B)** The total number of days that each intervention was used in our dataset, aggregated across regions but separated by country. The dashed vertical line indicates the total-number of region-days in our dataset. **(C)** Additional example timelines showing the cases, deaths, and interventions in 6 regions. Comparing two regions within England (Lincolnshire and Greater Manchester S.W.) and within Switzerland (Zürich and Géneve) reveals significant subnational variation, both in the interventions used and in the evolution of the epidemic.

Using this modelling approach, we estimate intervention effect sizes and express these as percentage reductions in the (instantaneous) reproduction number *R*_*t*_ (Fig. 2). The effect sizes in the second wave were considerably smaller than those estimated for the first half of 2020. All NPIs included in the study together reduced *R*_*t*_ by 66% [95% CI: 61%-69%], compared to medianreductions of 77%-82% in the first wave (*1, 2*). The difference between the waves is even more pronounced if we consider the effectiveness of the most stringent set of NPIs actually implemented in each region, rather than the (hypothetical) combined effectiveness of all NPIs included in the study. The most stringent set of NPIs implemented in each region reduced *R*_*t*_ by an average of 56% [95% CI: 40%-64%], compared to 76%-82% in the first wave, even though NPIs in the second wave were often similarly strict or stricter (*1, 2*). Finally, *R*_*t*_ was reduced from an average maximum of 1.7 [95% CI: 1.4-2.4] to a minimum of 0.7 [95% CI: 0.5-0.8] across regions in the second wave, compared to an average maximum of 3.3-3.8 and minimum of 0.7-0.8 in the first wave (*1, 2*).

**Fig. 2.**
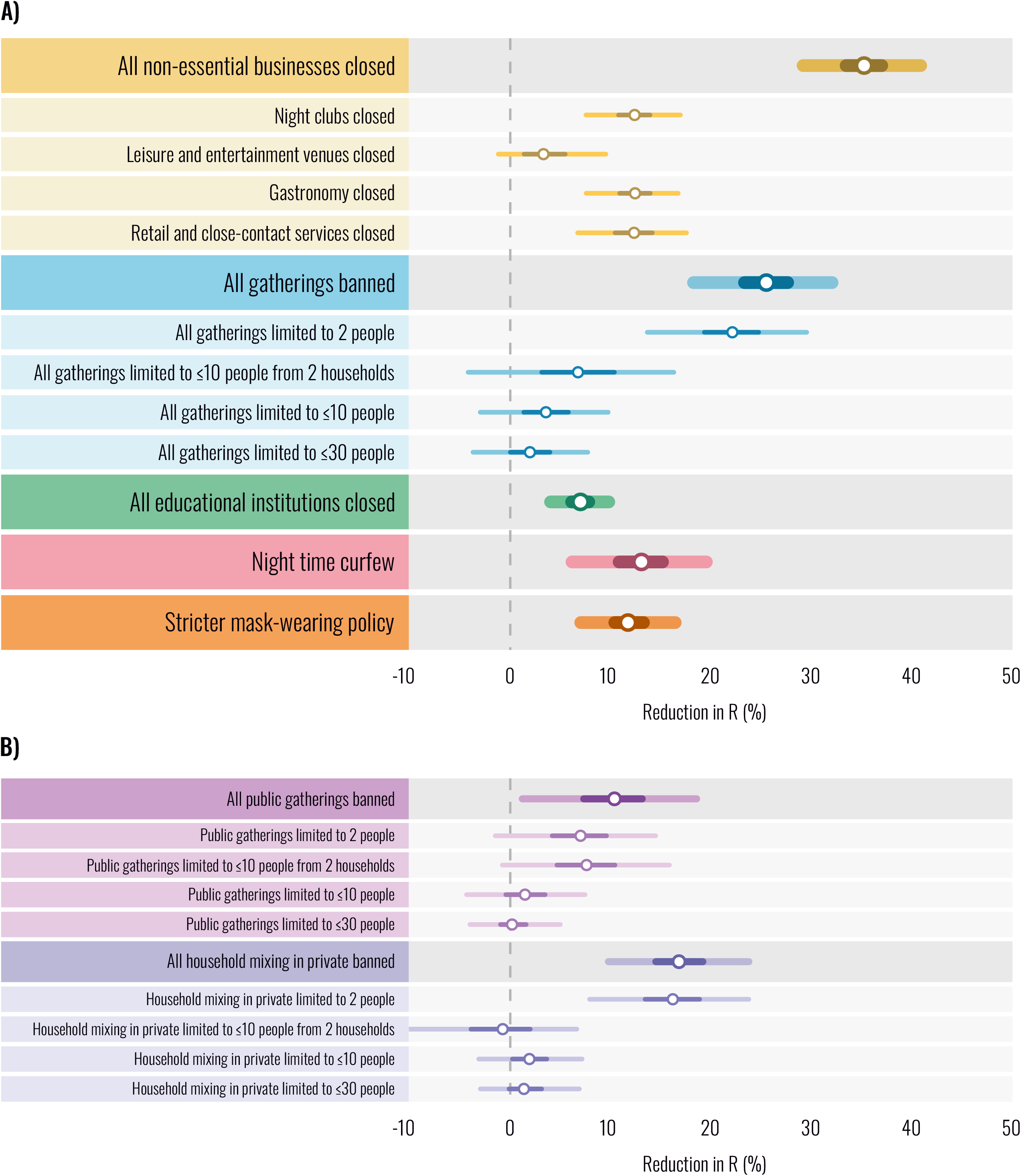
Intervention effectiveness under default model settings. Posterior percentage reductions in *R*_*t*_. Markers indicate posterior median estimates. Lines indicate the 50% and 95% posterior credible intervals. A negative 1% reduction refers to a 1% increase in *R*_*t*_. **(A)** Effectiveness of the main interventions included in our study. Intervention names preceded by “All” show the combined effect of multiple interventions. For example, “All gatherings banned” shows the combined effect of banning all public gatherings and all household mixing in private. **(B)** Individual effectiveness estimates for gathering types, separated into public gatherings and household mixing in private.

The most important explanation for these differences between the first and second wave is likely a combination of persistent behavioural change, such as avoiding close contact or increased hygiene, and the widespread adoption of safety protocols, such as distancing rules and adequate ventilation. These changes likely made various areas of public life safer and thereby reduced the effect of strict bans or closures. This hypothesis is corroborated by the stability of behavioural polls after the first wave (*1*). Several other factors seem less important but may still have contributed to the variation. First, a build-up of population immunity likely does not explain the reduction in NPI effectiveness: attack rates were low in our period of analysis (*1*) and the in-or exclusion of population immunity did not change the estimated effect sizes. Second, reduced adherence to NPIs in the second wave may have played a role, although adherence seems much more relevant for some NPIs (e.g. restrictions on private gatherings) than for others (e.g. closure of businesses or educational institutions). Finally, the ascertainment rate of cases increased (*30, 31*) throughout the first wave but was far more stable in the second wave. However, this difference is expected to increase the estimated effectiveness of NPIs in the second wave compared to the first, the opposite of what we found.

If all NPIs together reduced the original value of *R*_*t*_ to 18-23% in the first wave and to 35% in the second wave, this suggests an overall reduction in *R*_*t*_ of 34-49% due to behaviour changes, safety protocols, and potentially other effects. The result underscores the importance of viewing the effectiveness of an NPI relative to the counterfactual safety measures and behaviour in the absence of the NPI. Since first-wave NPI effectiveness was measured relative to pre-pandemic behaviour, it is likely inadequate to inform policy during an ongoing pandemic. While behaviour will continue to change dynamically, the increased stability seen in behavioural polls (*1*) means that second wave NPI effectiveness estimates are expected to generalise better to future waves.

### A detailed and subnational assessment of interventions in Europe’s second wave

We find that business closures were particularly effective, with a combined effect of reducing *R*_*t*_ by 35% [95% CI: 29-41%] (Fig. 2A). Closures of gastronomy (restaurants, pubs, and cafes) had a large effect on transmission with an estimated reduction in *R*_*t*_ of 12% [95% CI: 8-17%], an estimate that is broadly in line with the increases estimated to have occurred as a consequence of the UK’s “eat out to help out” scheme (*1*) in August. We find a similar substantial effect for closing night clubs [12%, 95% CI: 8-17%], which were predominantly shut earlier than other businesses; this effect size may reflect early second wave superspreading (*1*). The combined effect of closing retail and close contact services (such as hairdressers and beauty salons) is also considerable [12%, 95% CI: 7-18%]. Assuming that much of this effect is due to the more common type of business – namely, retail – it underscores the potential risks of brief but very numerous indoor contacts (*1*). Closing leisure and entertainment venues such as zoos, museums, and theatres had a small effect [3%, -1-10%].

As a broad intervention, we found that banning all gatherings, including 1-on-1 meetings, had a large effect: a 26% [95% CI: 18-32%] reduction in *R*_*t*_. By recording the number of persons and households allowed to meet, we were able to test the impact of various thresholds. We found no evidence for diminishing returns in the number of persons allowed to meet; indeed, only strict thresholds had a statistically significant effect. This is consistent with previous studies on the English tier 1 and tier 2 restrictions, which limited gatherings to 6 people and had a small total impact in contrast to the more stringent tier 3 (*18, 19, 35*). The small effect associated with more lenient person limits (10 or higher) contrasts with estimates from the first wave (*1*). The difference could be due to voluntary protective behaviours such as avoiding crowds and distancing, but also due to limited adherence to rules on private mixing (*1*). These factors may also explain why all gathering bans combined were somewhat less effective compared to business closures. Defining a “lockdown” policy as a ban on all gatherings and closure of all non-essential businesses, we estimate a total reduction in *R*_*t*_ of 52% [95% CI: 47-56%].

Most countries adopted different limits for public gatherings and household mixing in private at various times. We are therefore able to disaggregate the effect of these gathering types and examine their relative effects (Fig. 2B). While the total effect of banning all private mixing exceeds that of banning public gatherings, private mixing restriction was only effective at a strict threshold of 2 people allowed to meet. As discussed above, this could be due to a combination of low adherence and individuals voluntarily avoiding crowds.

Observational studies of the first wave consistently found that closing educational institutions was among the most effective NPIs (*2, 11*–*13, 37*). In strong contrast, we discover that their effect was small in the second wave [7%, 95% CI: 4-10%]. We conjecture that a combination of safety measures, behaviour changes, and epidemiological factors (*1*) in the education sector prevented large undetected clusters which may have developed in the first half of 2020 (*1*–*1*). Schools in Europe’s second wave operated under safety measures that some other organizations lacked, including symptom screening, asymptomatic testing, contact tracing, sanitizing, ventilation, distancing, reducing group sizes, and preventing the mixing of groups (*39, 43*). However, measures can vary by country (*1*) and detailed further assessments are required. Without sufficient measures, opening schools could lead to a resurgence (*1*).

We documented student presence separately for universities (or higher education) and schools (both primary and secondary) by recording their local term times, holidays, and closure dates in all 114 regions, and identifying regions without universities. However, we refrain from quantifying the relative effects of schools and universities here, and instead only report their combined effect since it was more robust than the individual effects in one sensitivity analysis that was designed to adjust for undetected infections (*1*) in schools (Supplementary text 1.1.3). The small combined effect suggests that safety measures and behaviour changes significantly reduced transmission associated both with schools and with universities.

Finally, we find that a stricter mask-wearing policy (mandatory in most or all shared/public spaces) and a night time curfew had moderate, but statistically significant effects [12%, 95% CI: 7-17%] and [13%, 6-20%]). However, due to the broad nature of these interventions, they are also likely to interact with other active NPIs. In contrast, the other NPIs affect largely distinct areas of social activity and therefore are not expected to mutually interact to a great extent.

### Robustness of estimates

The usefulness of estimates of intervention effectiveness hinges on their robustness, and sensitivity analysis can help diagnose when the model is misspecified or the data too collinear (*27, 46*). Fig. 3 shows how the median estimates of effect size from Fig. 2 vary across 17 types of sensitivity analyses, as we modify the priors and structure of the model, change the distributions of epidemiological delays, and randomly vary the set of regions and other aspects of the data. Each analysis is shown in Supplementary text section 1.1. Since we cannot model all possible factors that affect transmission, we also investigate the stability of effects sizes to unobserved effects that influence *R*_*t*_, acting as possible confounders. These include unrecorded NPIs and changes to ascertainment and fatality rates. See Supplementary Table S1 for details on the experiments in Fig. 3 and Supplementary text sections 1.2-1.6 for additional validation experiments including simulations on confounding, posterior predictive checks (*1*), and a single-model meta-analysis across regions.

**Fig. 3.**
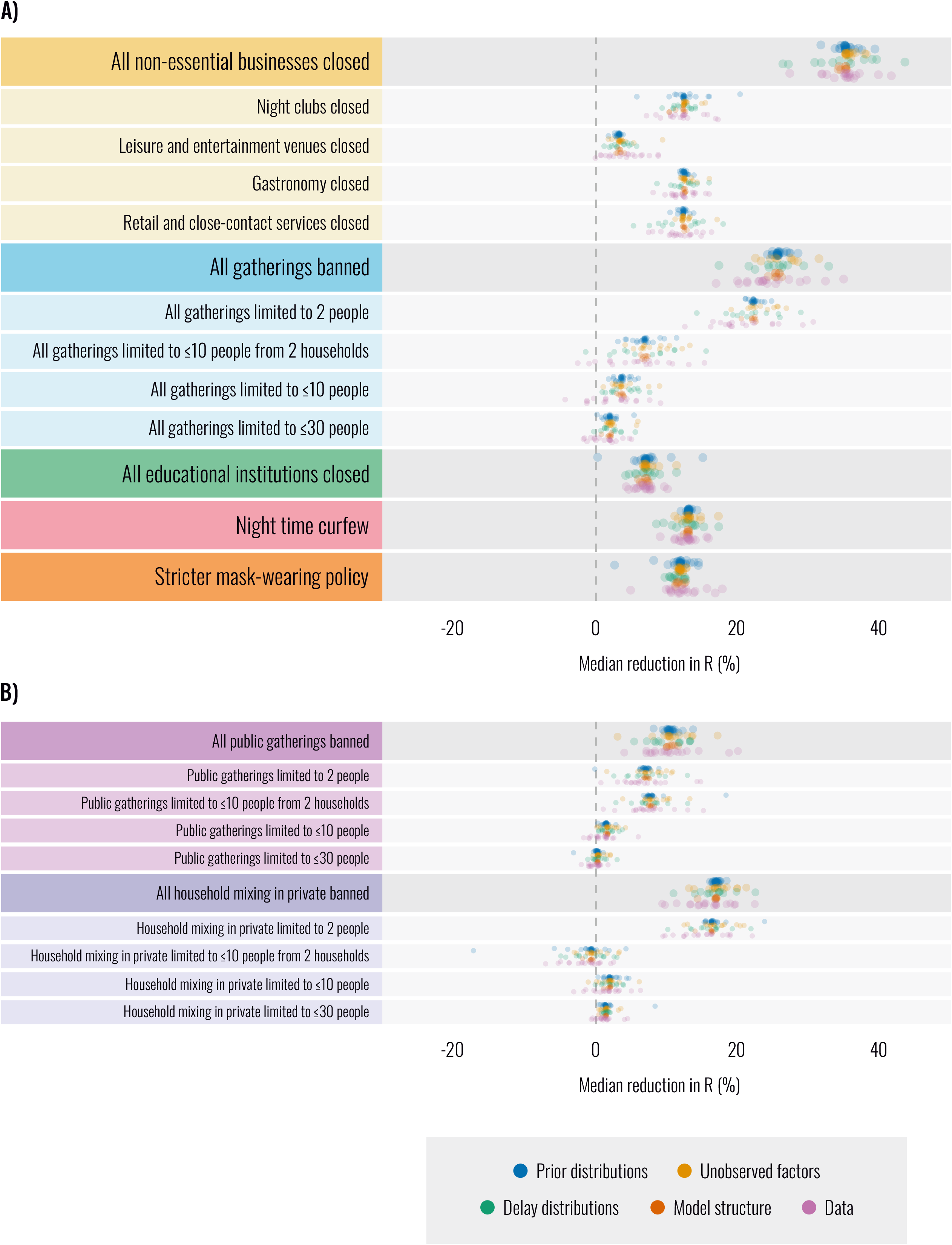
Robustness of median intervention effectiveness estimates across 86 experimental conditions. Each dot represents the posterior median intervention effectiveness in a different experimental condition. Dot color indicates five categories of sensitivity analyses. Each category contains several sensitivity analyses (17 in total). Each sensitivity analysis contains several experimental conditions (86 in total). Supplementary Table S1 lists all sensitivity analyses by category. **(A)** Robustness of effectiveness estimates of the main interventions included in our study. Intervention names preceded by “All” show the combined effect of multiple interventions. For example, “All gatherings banned” shows the combined effect of banning all public gatherings and all household mixing in private. **(B)** Robustness of the individual effectiveness estimates for separately banning public gatherings or household mixing in private.

Median effects vary across a total of 86 experimental conditions in Fig. 3. However, a robust broad picture emerges in which some NPIs outperform others across all experiments. Although our results are robust to varying strengths and types of unobserved factors, the true strength of unobserved confounding is unknown and our study is therefore subject to the limitations of observational approaches.

## Conclusion

European governments are presently deciding which interventions to keep and which to remove. These decisions demand effectiveness estimates directly from the second wave since estimates from the first wave are measured relative to pre-pandemic contact patterns, and therefore overestimate NPI effectiveness in an ongoing pandemic.

As different areas of social life reopen, effectiveness estimates can inform which areas require additional safety measures to reopen safely. In case of a resurgence--for example, due to variants of concern that are more transmissible or evade protection from previously acquired immunity--our results provide a starting point to effectively control infections while minimizing social and economic harms.

Using a European NPI dataset of increased scale and granularity enabled us to provide the first effectiveness estimates for individual NPIs in Europe’s second wave. However, decision-makers using these results should account for several factors. Discontinuing safety measures and voluntary protective behaviours among the still-susceptible population would change intervention effectiveness, perhaps even increasing it to first-wave levels. Mass vaccination may increase the relative importance of interventions such as school closures that mainly affect the unvaccinated population. Moreover, it is still unknown whether variants of concern differentially affect certain population groups. Finally, while our model allows for spatiotemporal variation in behaviour and adherence, no single effectiveness estimate can apply to all regions. Therefore, our estimates should be combined with expert judgment to adjust them to local and contemporary circumstances

## Methods

### Data

#### Dataset overview

We collected a custom NPI dataset for this modelling study, as existing datasets do not provide sufficient geographical resolution to model the second wave. Further advantages of our dataset are NPI definitions tailored towards the second wave and high data quality through extensive validation.

To create this dataset, we collected chronological data on NPIs that were in place between 1 August 2020 and 9 January 2021 in administrative regions, districts, and local areas of 7 European countries. The resulting dataset contains over 5,500 entries on various NPIs in 114 regions of analysis (Supplementary Table S4). Every entry includes the NPI start date and end date, quotes and comments, and one or more sources from websites of governments and universities, legal documents, and/or media reports. Daily case and death data were obtained from government websites (Supplementary Table S3). The data is available upon request.

We now describe how we selected the countries, regions of analysis within each country, and NPI definitions.

We first identified 7 European countries for which public data on daily reported cases and deaths were available at the same geographical resolution at which the country implemented NPIs (Austria, the Czech Republic, England, Germany, Italy, the Netherlands, Switzerland).

To gather initial information about the transmission-reducing NPIs used in these countries, we conducted an exploratory data collection and interviewed local epidemiologists from the countries. Based on these data, we created NPI definitions that faithfully represent the interventions that were implemented in these countries. We focused on clear-cut, major interventions that were implemented in many countries and we only recorded mandatory restrictions, not recommendations^1^. We also accounted for closures that are not due to NPIs, such as vacation and term times in schools and universities, as we surmised that these effectively function as NPIs.

The exploratory data also informed the appropriate level of geographical granularity for the NPI data collection. In each country, we set our regions of analysis to correspond to the highest possible level of administrative division for which NPI implementations were identical throughout each region. The chosen administrative divisions were (Supplementary Table S3):

- states in Austria,
- administrative regions in the Czech Republic,
- NUTS 3 statistical regions in England,
- districts in Germany,
- administrative regions in Italy^2^,
- safety regions in the Netherlands,
- and cantons in Switzerland.

For Austria, the Czech Republic, Italy, and the Netherlands, it was feasible to collect data from the whole country (9, 14, 21, and 25 regions of analysis). From each other country, we took a stratified random sample of 15 regions of analysis^3^. The sample was stratified by the regions’ number of COVID-deaths in the first wave, to ensure a sufficiently diverse sample and reduce the variance of our NPI effect estimator. To increase statistical power and reduce the influence of imported infections, we did not include regions with fewer than 2000 reported cases during the analysis period.

#### Data collection

To ensure high data quality, the NPI data was collected with semi-independent double entry and several validation steps. Each country was collected by two authors of this paper, who were provided with a detailed description of the NPIs. The researchers manually researched all dates by using internet searches and screening (local) government press releases, ordinances, and legislation. There was no automatic component in the data gathering process.

In the first round of data entry, the researchers initially collected the timeline of national NPI implementations. The researchers then compared their national timeline to the Oxford COVID-19 Government Response Tracker dataset (*1*) and, if there were any conflicts, visited all primary sources to resolve them. The data for each region of analysis were then entered by one of the two researchers, drawing on the national timeline and additional research. Several countries operated a tier or trafic light system that governed NPI implementation in subnational administrative divisions. For these countries, the researchers did not blindly enter the NPIs prescribed by the tier or traffic light system, but additionally consulted local government websites and media reports to investigate if the NPIs prescribed by the national system were, in fact, implemented in a region of analysis.

In the second round of data entry, every entry was independently entered again by another researcher. This researcher had access to the sources found in the first round as well as the associated quotes and comments, but not to the NPI data entered in the first round. This semi-independent double entry is similar to the validation used for parts of CoronaNet (*1*).

Finally, data from the two rounds of entry were compared and all conflicts resolved by discussion and by visiting primary sources. A researcher then manually compared the data from all countries to ensure the consistent application of NPI definitions across countries. We also validated the data against further external sources (e.g. (*1*) for Italy or (*1*) for England), contacted local epidemiologists when in doubt, and implemented a range of automated plausibility checks.

Throughout the data collection process, the researchers discussed edge cases and judgement calls on a shared online workspace, to ensure consistency across countries. As expected, the validation process removed various sources of error and inconsistency. Supplementary text section 4 contains detailed explanations on coding decisions and judgement calls.

The total time spent on manual data collection, not including the design of the process, was 950 hours, with 185 hours on the national timelines, 470 hours on collecting the regions of analysis, and 290 hours on the validation steps.

#### Data pre-processing

To prevent bias, we excluded^4^ all observations in a region of analysis after the date when the variant of concern (VOC) B.1.1.7 first made up >10 % of infections in that region. Specifically, we excluded cases 5 days after >10 % of all infections were due to the VOC and deaths 11 days after this value was reached. We chose these values to ensure that on the last included day, more than 80% of the reported cases and deaths were generated **before** the VOC exceeded >10 % of all infections, according to our delay distributions. This only affected regions in England, usually towards the end of November (*1*).

The last day recorded in the intervention data set is 9th January 2021. Therefore, we included cases up to 5 days after this date and deaths up to 11 days after this date, as they are predominantly generated by infections before the 9th of January (see above).

Furthermore, to prevent influence from infections generated before the start of the analysis period, we excluded cases in the first 8 days (until 8th August) and deaths in the first 25 days (until 25 August). These values were chosen such that 80% of the cases and deaths recorded on the first observed day were generated in our window of analysis (including seeded infections), according to our delay distributions.

Supplementary text section 3.1 explains how we created the NPI features used in the modelling from the raw data. The final NPIs used for modelling are described in Table 2.

**Table 1.** Main dataset characteristics. In total, we collated more than 5,500 intervention entries through a bespoke systematic categorisation.

|  |  |
| --- | --- |
| Countries | 7 |
| Regions of analysis | 114 |
| Period | 1 August 2020 - 9 January 2021* |
| Days across all regions | 19,000 |
| NPI entries in the dataset | > 5,500 |
| Data validation (manual) | Semi-independent double entry;<br>interviews with local epidemiologists;<br>validation against external sources;<br>cross-country consistency checks |

\*We ended the period of analysis before 9 January 2021 for English regions depending on their prevalence of a new variant of concern (see *Methods*).

**Table 2.**
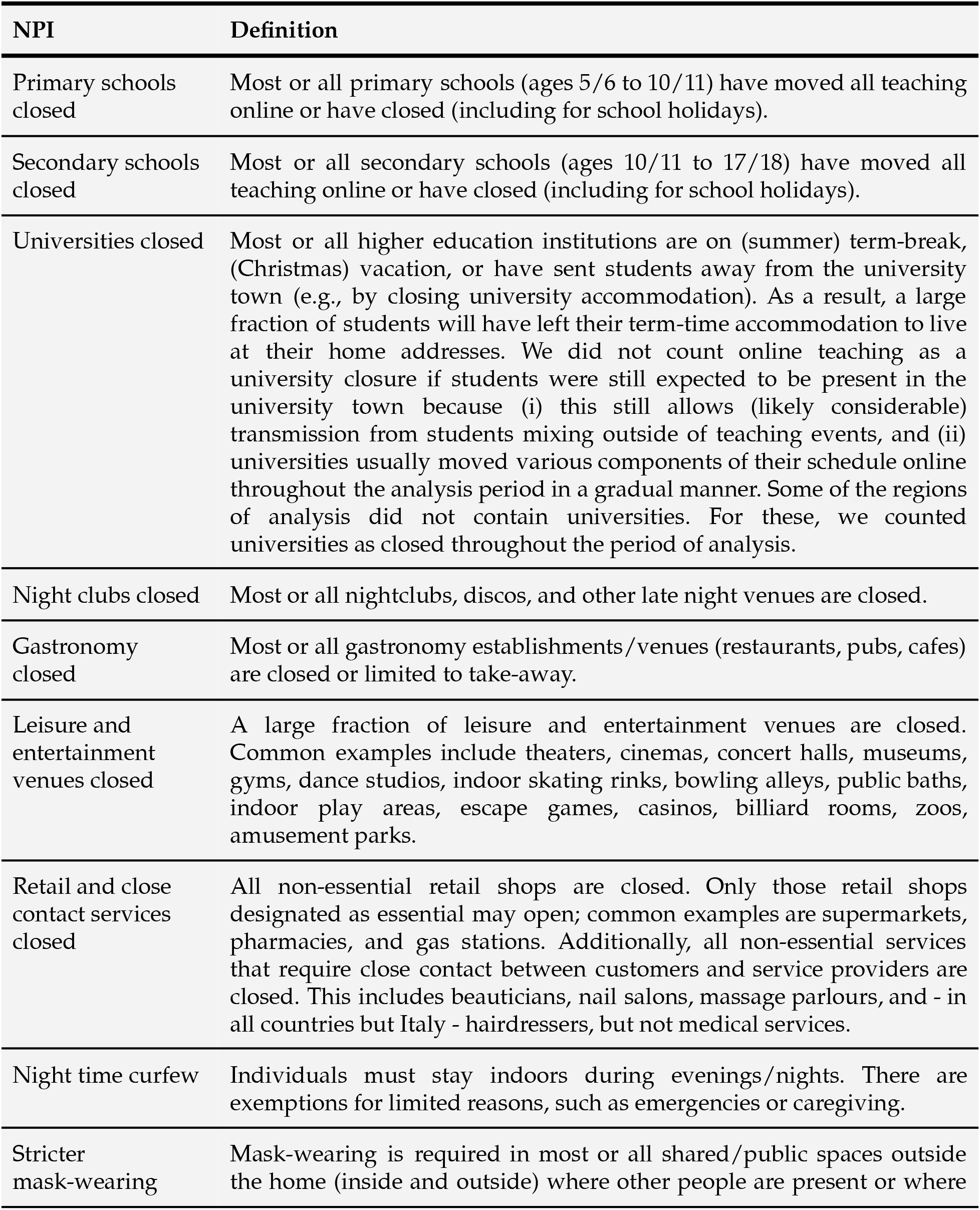
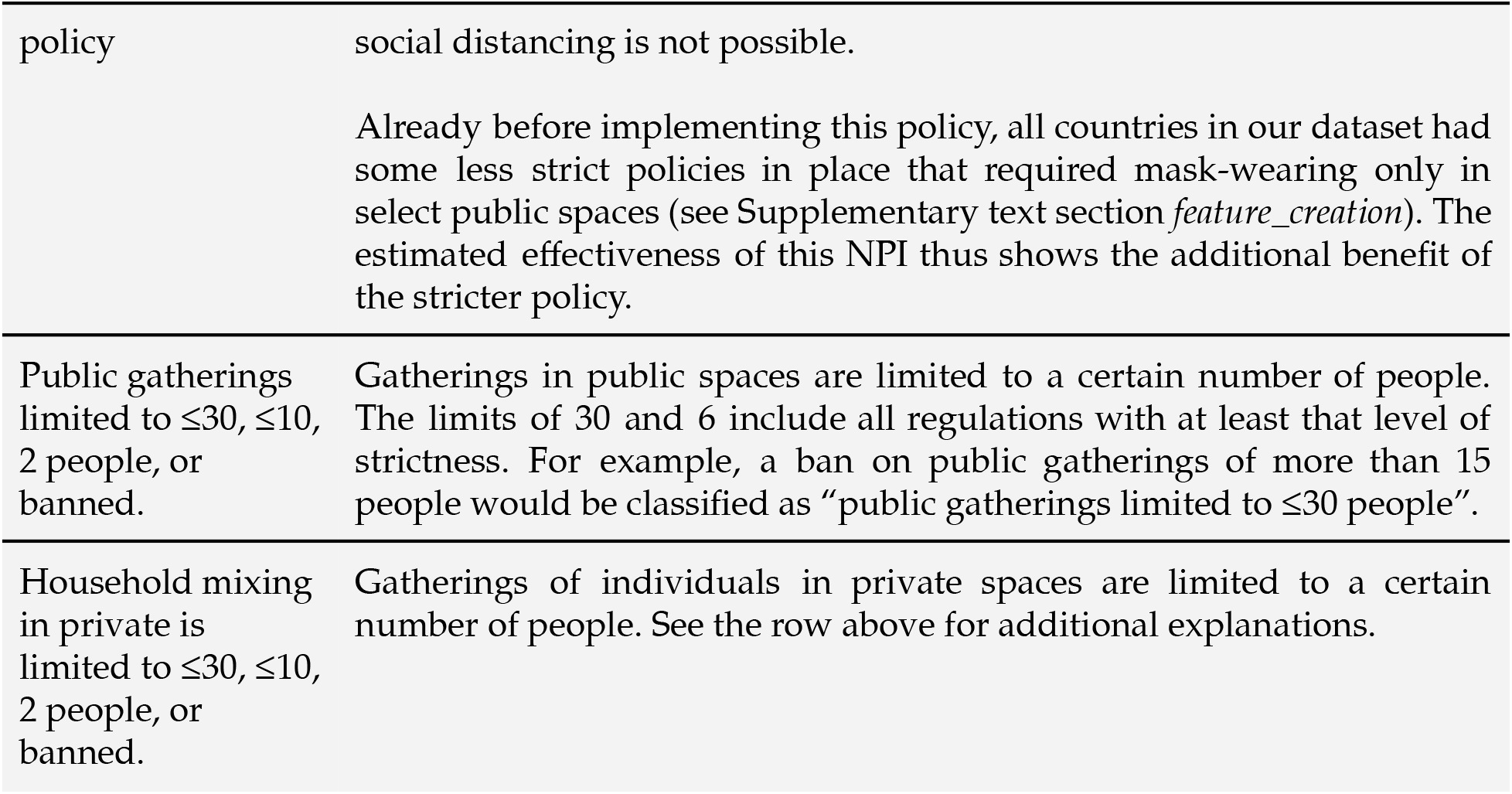
NPI definitions.

### Model

Please refer to the supplement for the model description.

## Data Availability

Data available upon request

## Acknowledgements

We thank Fabian Valka for advice on the Austrian COVID response, Ilaria Dorigatti for advice on Italy, Natalie Claire Ceperley for advice on Switzerland, Veronika Nyvltova for help with Czech NPI data, Paul Hunter for sharing his UK tier data, Toby Philips for advice on NPIs implemented in the second wave. We thank Muhammed Razzak for his comments on the manuscript.

M. Sharma was supported by the EPSRC Centre for Doctoral Training in Autonomous Intelligent Machines and Systems (EP/S024050/1) and a grant from the EA Funds programme. S. Mindermann’s funding for graduate studies was from Oxford University and DeepMind. C. Rogers-Smith was supported by a grant from Open Philanthropy. A.J. Norman was supported by the U.K. BBSRC and Open Philanthropy. J Ahuja was supported by Open Philanthropy. J. T. Monrad was supported by the Augustinus Foundation, the Knud Højgaard Foundation, the William Demant Foundation, the Kai Lange and Gunhild Kai Lange Foundation, and the Aage and Johanne Louis-Hansen Foundation. G.Leech was supported by the UKRI Centre for Doctoral Training in Interactive Artificial Intelligence (EP/S022937/1). S.B. Oehm was supported by the Boehringer Ingelheim Fonds. L.Chindelevitch and S.Bhatt acknowledge funding from the MRC Centre for Global Infectious Disease Analysis (MR/R015600/1), jointly funded by the U.K. Medical Research Council (MRC) and the U.K. Foreign, Commonwealth and Development Office (FCDO), under the MRC/FCDO Concordat agreement; are part of the EDCTP2 program supported by the European Union; and acknowledge funding by Community Jameel. S. Flaxman acknowledges the EPSRC (EP/V002910/1) and the Imperial College COVID-19 Research Fund. J.M. Brauner was supported by the EPSRC Centre for Doctoral Training in Autonomous Intelligent Machines and Systems (EP/S024050/1) and by Cancer Research UK. S. Bhatt acknowledges The UK Research and Innovation (MR/V038109/1), the Academy of Medical Sciences Springboard Award (SBF004/1080), The MRC (MR/R015600/1), The BMGF (OPP1197730), Imperial College Healthcare NHS Trust-BRC Funding (RDA02), The Novo Nordisk Young Investigator Award (NNF20OC0059309) and The NIHR Health Protection Research Unit in Modelling Methodology. S. Bhatt thanks Microsoft AI for Health and Amazon AWS for computational credit

## Author contributions

S. Mindermann, M. Sharma, J.M. Brauner, S. Mishra, S. Bhatt, Y. Gal, L. Chindelevitch, S. Flaxman conceived the research.

J.M. Brauner, S. Mindermann, M. Sharma, S. Bhatt, S. Mishra, B. Snodin, G. Altman, J. Ahuja, L. Finnveden, J.B. Sandbrink, G. Dhaliwal, J.T. Monrad, A.J. Norman, J, Kulveit, G. Leech, J.F. Sandkühler designed and conducted the NPI data collection.

M. Sharma, S.Mindermann, S. Mishra, J.M. Brauner, S.Bhatt, Y. Gal, G. Leech, S. Flaxman, L. Chindelevitch designed the model and modelling experiments.

M. Sharma, S. Mishra, C. Rogers-Smith, G. Leech, L.Finnveden performed and analyzed the modelling experiments.

S. Mindermann, S. Mishra, J.M. Brauner, S. Bhatt, J.T. Monrad, J. Kulveit did the literature review.

S. Mindermann, S. Bhatt, J.M. Brauner, M. Sharma, S. Mishra, J.T. Monrad, S.B. Oehm, G. Leech, A.J. Norman, C. Rogers-Smith, B. Snodin, G. Dhaliwal, G. Altman, J. Ahuja, J.B. Sandbrink wrote the manuscript.

All authors read, gave input on, and approved the final manuscript.

## Footnotes

1 Some regions do not have the legislative capacity to regulate gatherings in private homes. As an exception, we therefore also recorded prohibitions against private gatherings that were not legally binding. This was applicable in Italy, Austria, and the German state Northrhine-Westphalia. In each case, we confirmed with locals that these restrictions were followed by the vast majority of the population.

2 With the exception of the Trentino-Alto Adige region, which was split into the autonomous provinces Trentino and Alto-Adige.

3 Each of the 16 German states had different regulations for its districts. To reduce the work required for data collection, we sampled the 15 districts only from the four largest states (Northrhine-Westphalia, Bavaria, Baden-Württemberg, Lower Saxony). These four states make up 60% of the population.

4 The observations were excluded by *masking* the observed cases and deaths, equivalent to removing these days from the likelihood/log-probability term. This ensures that these observations do not provide evidence, in the Bayesian sense, about our estimated NPI effects.

