## Supplement for "Understanding the effectiveness of government interventions in Europe’s second wave of COVID-19"

### 1. Methods

#### 1.1. Model

We construct a semi-mechanistic Bayesian hierarchical model, similar to that of Brauner et al. (*1*, *2*), but with adaptations tailored for the second wave. Namely, we allow for changes in transmission unrelated to NPIs, which allow the model to explain e.g., unrecorded interventions. Furthermore, we account for the variance inherent in low incidence settings. Our model implementation is available on GitHub (https://github.com/MrinankSharma/COVID19NPISecondWave).

We proceed by describing the model in Figure 1 from bottom to top.

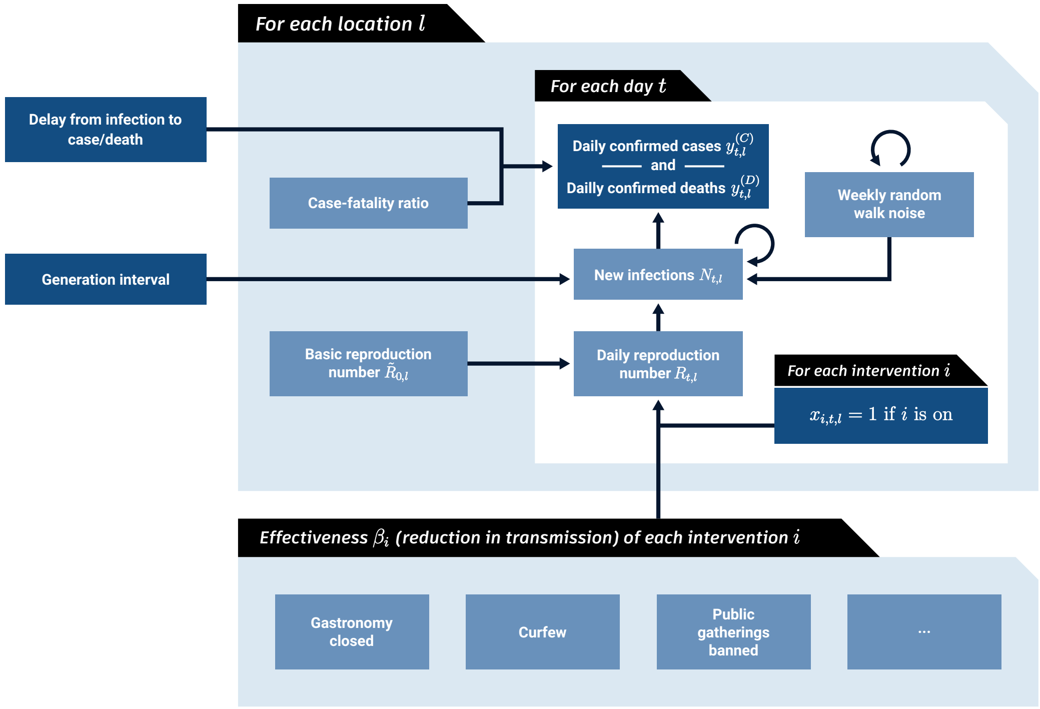

**Figure 1: Model Overview. Dark blue nodes are observed. We describe the diagram from bottom to top. The mean effect parameter of NPI $i$ is $\beta_i$. On each day $t$, a location's reproduction number $R_{t,l}$ depends on the basic reproduction number $\tilde{R}_{0,l}$, the NPIs active in that location and a location-specific latent weekly random walk. The active NPIs are encoded by $x_{i,t,l}$, which is 1 if NPI $i$ is active in location $l$ at time $t$, and 0 otherwise. A random walk flexibly accounts for trends in transmission due to unobserved factors. $R_{t,l}$ is used to compute daily infections $N_{t,l}$ given the generation interval distribution and the infections on previous days. Finally, the expected number of daily confirmed cases $y_{t,l}^{(C)}$ and deaths $y_{t,l}^{(D)}$ are computed using discrete convolutions of $N_{t,l}$ with the relevant delay distributions.**

*Reproduction number.* The epidemic's growth is described by the time-and-location-specific (instantaneous) reproduction number $R_{t,l}$. $R_{t,l}$ is the expected number of secondary infections that would arise from a primary infection at time $t$ in location $l$, provided conditions remain the same after time $t$. We allow $R_{t,l}$ to change over time, even if the interventions implemented in location $l$ do not change. In particular, the value of $R_{t,l}$ depends on three factors: a) the reproduction number at the start of the period in the absence of NPIs, $\tilde{R}_{0,l}$; b) the active nonpharmaceutical interventions (and their effectiveness); and c) a latent (weekly) random walk. The random walk term allows $R_{t,l}$ to change from one week to the next. Precisely, $R_{t,l}$ follows:

$$R_{t,l} = \underbrace{\tilde{R}_{0,l}}_{\substack{R \text{ at } t=0 \text{ if no} \\ \text{NPIs active}}} \overbrace{\left( \prod_{i=1}^{I} \exp\left(-\beta_i \, x_{i,t,l}\right) \right)}^{\text{effect due to active NPIs}} \underbrace{\exp(z_{t,l})}_{\substack{\text{latent random} \\ \text{walk}}} , \tag{1}$$

where $x_{i,t,l} = 1$ means NPI $i$ is active in location $l$ on day $t$ ($x_{i,t,l} = 0$ otherwise), and $I$ is the number of NPIs. We now explain each of these terms in more detail.

We place a prior distribution over $\tilde{R}_{0,l}$, the reproduction number (in the absence of NPIs) on August 1st 2020. In fact, many locations had some recorded interventions active at $t = 0$. Therefore, we chose the mean of the prior on $\tilde{R}_{0,l}$ carefully. We ensured the prior on $R_{0,l}$ matched published estimates[a] of $R_t$ for the first week of August from (*3*) and (*4*). For clarity, $\tilde{R}_{0,l}$ is the reproduction number that *would* have been observed in location $l$ at $t = 0$ had no NPIs been active. The prior over $\tilde{R}_{0,l}$ follows:

$$\tilde{R}_{0,l} \sim \text{Truncated Normal}(1.35, 0.3^2), \tag{2}$$

where trunction prevents values of $\tilde{R}_{0,l}$ less than 0.1.

We parameterise the effect of NPI $i$ with the effect parameter $\beta_i$. This parameter is independent of time and shared across all locations, i.e., the effectiveness of a particular NPI is assumed to be identical across regions (though the random walk described below can account for differences). We place an Asymmetric Laplace prior over the effect parameter $\beta_i$, with scale parameter 30, asymmetry parameter 0.5, and location parameter 0. This prior has mean 0.05 and standard deviation 0.07. The prior allows for (unbounded) positive and

[a] The prior over $R_{0,l}$ depends on $\tilde{R}_{0,l}$, the interventions active at $t = 0$ in location $l$ and the prior on the effectiveness of NPIs. We fixed the intervention prior first and then chose the prior on $\tilde{R}_{0,l}$

negative effects as we cannot exclude the possibility that an NPI increases transmission. However, our prior places 80% of its mass on positive effects, reflecting a belief that NPIs are more likely to reduce transmission than to increase it. Furthermore, this is a shrinkage prior–it places more than 80% of its mass on "small" effectiveness (less than 10% change in $R_{t,l}$).

The final component used to calculate $R_{t,l}$ is a location-specific latent random walk. This random walk allows for changes in $R_{t,l}$ every week that are due to factors outside the model. A random walk can explain *lasting* changes in transmission, unlike typical noise models. For example, suppose there was an unrecorded intervention in location $l$ at time $t$, or a recorded intervention with unusually low adherence. Then the random walk could be used to explain the observed change in transmission. Mathematically, the random walk noise terms follow:

$$z_{t,l} = \begin{cases} 0 & t \leq 13 \\ z_{t-1,l} + \varepsilon_{\lfloor (t-14)/7 \rfloor, l} & \text{if } t \bmod 7 = 0 \,, \\ z_{t-1,l} & \text{otherwise} \end{cases} \tag{3}$$

where $\lfloor \cdot \rfloor$ denotes the floor operation and $\epsilon_{i,l} \sim \text{Normal}(0, \sigma_R^2)$. In words, $z_{t,l}$ is set to 0 for the first two weeks, meaning that $R_{t,l}$ depends only on $\tilde{R}_{0,l}$ and the active interventions for the first two weeks. Then, every week, the value of $z_{t,l}$ may increase or decrease depending on the noise variable $\epsilon_{i,l}$. If we observe that transmission increased in a particular week, then we may infer $\epsilon_{i,l}$>0 and vice versa.

The random walk addresses an important limitation–we cannot include all possible factors that affect transmission. We can attempt to attribute effect sizes to NPIs at a time $t$, but we need to agnostically account for other unobserved factors that could have changed transmission (e.g. behaviour, adherence). By using a random walk, we include a latent stochastic process that agnostically models unobserved trends and residual structural correlations.

Furthermore, we place a prior over $\sigma_R$, which describes the scale of the random walk process. As $\sigma_R$ increases, the latent random walk can be used to explain larger changes in transmission. An advantage of placing a prior over $\sigma_R$ and performing joint Bayesian inference is that, if warranted by the data, an appropriate value may be inferred automatically. Our prior is $\sigma_R \sim$ Half Normal(0.15). We include this prior distribution in our sensitivity analysis (Fig. S10) and find low sensitivity. Furthermore, we find that the data provide strong evidence about the value of $\sigma_R$ (see Fig. S30 for a posterior and prior comparison).

*Infection process.* Let $N_{t,l}$ denote the number of new infections at time $t$ in location $l$. Furthermore, the generation interval (GI), which is the time between successive infections in a transmission chain, is denoted with the distribution $\pi_{GI}[\tau]$ where $\tau$ refers to the number of days since infection. The expected number of infections then follows a discrete renewal process (*2,5*):

$$\bar{N}_{t,l} = R_{t,l} \sum_{\tau=1}^{32} \left( \bar{N}_{t-\tau,l} \cdot \pi_{GI}[\tau] \right). \tag{4}$$

Renewal processes have a strong relationship to Hawkes processes and arise naturally from a Bellman Harris branching process (*6,5*). The renewal equation has also been shown to be equivalent to a susceptible-exposed-infected-recovered Erlang model. The renewal equation therefore specifies an epidemilogically motivated function class. One issue with the renewal equation is that it specifies a deterministic expectation for the number of new infections. This is generally suitable as infections become large, but in low incidence settings, estimation of $R_{t,l}$ can be sensitive to random fluctations and noise. Therefore, we include an additive noise term, reflecting a belief that changes in the number of infections at low infection counts provide limited evidence to ascertain $R_{t,l}$, and must be treated with caution. Thus, the actual number of infections follows:

$$N_{t,l} = \text{softplus}(\bar{N}_{t,l} + \epsilon_{t,l}), \tag{5}$$

where $\epsilon_{t,l}^{(N)} \sim \text{Normal}(0, 5^2)$ (sensitivity analysis in Fig. S9). We use the softplus$(\cdot)$ rectifier to ensure that $N_{t,l} \geq 0$.

We seed the model with one week of unobserved initial infections[b].

$$N_{-t,l} = \text{Lognormal}(\tilde{\mu} = 0, \tilde{\sigma} = 3), \qquad \text{for } 1 \leq t \leq 7. \tag{6}$$

*Infection ascertainment and fatality rates.* Scaling all values of a time series by a constant maintains its reproduction numbers. Our model is thus invariant to the scale of the observations and therefore to *time-invariant* differences between locations in the Infection Fatality Rate (IFR), which is the proportion of infected people that subsequently die, and the Infection Ascertainment Rate (IAR), which is the proportion of infected people who are subsequently tested positive. Since the model is invariant to the absolute scale of these

[b]Since we treat new infections as a continuous number, their initial value can be between 0 and 1.

rates, we set $\text{IAR}_l = 1$ for all local areas, and we place a prior over $\text{IFR}_l$. Both the IAR and IFR are assumed to be constant over time. Additionally, since we assume $\text{IAR}_l = 1$, the IFR is actually a *case*-fatality rate and the variable $N_{t,l}$ effectively represents the infections that are later confirmed as positive cases. The uninformative prior over $\text{IFR}_l$ follows:

$$\text{IFR}_l \sim \text{Uniform}[10^{-3}, 1]. \tag{7}$$

We then have:

$$N^{(C)}_{t,l} = N_{t,l}, \text{ and } N^{(C)}_{t,l} = \text{IFR}_l \cdot N_{t,l}. \tag{8}$$

As such, $N^{(C)}_{t,l}$ represents infections that are later confirmed, and $N^{(D)}_{t,l}$ represents infections that later result in death.

As part of our validation, we replace the assumed time-constant IFR and IAR with their estimates in England (applying these to all countries), taken from (*7*). These time-varying estimates of the IFR/IAR are estimated using seroprevalence data from ONS (*8*) and REACT (*9*), along with case and death time series for England. See Figure S22. We find that our NPI effectiveness estimates are not sensitive to this change.

*Observation model for cases.* The expected number of confirmed cases on day $t$ in location $l$ is given by a discrete convolution:

$$\bar{y}^{(C)}_{t,l} = \sum_{\tau=0}^{31} N^{(C)}_{t-\tau,l} P_C(\text{delay} = \tau), \tag{9}$$

where $P_C(\text{delay})$ is the distribution of the delay from infection to case-reporting. This distribution is truncated to 31 days for computational efficiency. As in prior works (*10, 1*), the observed cases $y^{(C)}_{t,c}$ follow a negative binomial distribution with mean $\bar{y}^{(C)}_{t,c}$ and a country-specific inferred dispersion parameter, $\Psi^{(C)}_{c(l)}$. Since different countries have different reporting practices, we allow $\Psi^{(C)}_c$ to differ by country. The prior over this parameter is as follows:

$$\Psi^{(C)}_c \sim \text{Half Normal}(5). \tag{10}$$

*Observation model for deaths.* The expected number of deaths on day $t$ in location $l$ is given by a discrete convolution:

$$\bar{y}_{t,l}^{(D)} = \sum_{\tau=0}^{63} N_{t-\tau,l}^{(D)} P_D(\text{delay} = \tau), \tag{11}$$

where $P_D(\text{delay})$ is the distribution of the delay from infection to death reporting. Similar to cases, the delay vector is truncated for computational reasons, but since the delay between infection and death is longer, we truncate this distribution to a maximum delay of 63 days.

Finally, the observed deaths $y_{t,c}^{(D)}$ follow a negative binomial distribution with mean $\bar{y}_{t,c}^{(D)}$ and a country-specific inferred dispersion parameter, $\Psi_c^{(D)}$:

$$\Psi_c^{(D)} \sim \text{Half Normal}(5). \tag{12}$$

Having separate dispersion parameters for cases and deaths ensures that they can be weighted differently if there is a difference in their output variance.

The model was implemented in NumPyro (*11*). We infer the unobserved variables in our model using the No-U-Turn Sampler (NUTS) (*12*), a standard Markov chain Monte Carlo sampling algorithm. We used 4 chains with 250 warmup samples and 1250 draw samples, thereby obtaining 5000 posterior samples. We ensured that the posterior had converged by ensuring there were no divergence transitions, as well as monitoring the effective sample size and rank-normalized split-$\hat{R}$ statistic.

#### 1.2. Delay distributions

*Case and death delays.* Recall that our model requires external knowledge of the delay between infection and case confirmation as well as the delay between infection and death reporting. Many previous studies use estimates for delay distribution based on the data from the first wave (*1, 2, 13*). However, these delay distributions may be different in the second wave due to sustained investment in testing capabilities and healthcare. Therefore, we re-estimate these delay distributions using data from the second wave.

The delay from infection to case confirmation is composed of the *incubation period*–the time from infection to onset of symptoms–and the *symptom-to-confirmation* delay. Similarly, the delay from infection to death reporting is composed of the incubation period and the *symptom-to-death reporting* delay. We take an estimate of the incubation period from a meta-analysis (*14*). We then combine this incubation period with estimates of the symptom-to-confirmation delay and the symptom-to-death reporting delay from linelist data to form our total delay distributions.

We use linelist data from Austria, Germany and the United Kingdom (UK). This linelist data contains country-specific patient data of the date of symptom-onset, the date of case confirmation (for Austria, Germany and the UK) and the reported date of death (for Austria and the UK). To ensure that the linelist data we used was appropriate for the second wave, and to avoid censoring bias, we filtered the linelist data using the following conditions:

- date of onset of symptoms ≥ 2020/07/01
- date of onset of symptoms ≤ 2020/11/01
- date of death ≤ 2021/01/22
- date of death ≥ date onset
- date of admission ≥ date onset
- date of test confirmation ≥ date onset

By neglecting symptom onsets dates past November, we mitigate censoring bias. There were almost 3 months since November for the latest possible onset date to fully evolve. Furthermore, by filtering the date of admission to be after the symptom-onset date, we prevent bias from hospital-acquired infections.

We fitted gamma distributions to the onset-to-confirmation and onset-to-reported-death data. We also fit Weibull, Lognormal, and Negative Binomial distritbutions to the data but, using model selection (*15*), found these to have an inferior fit. The fitted gamma distribution for the onset-to-confirmation delay has mean 5.28 days and standard deviation 3.75 days. The fitted gamma distribution for the onset-to-reported-death delay has mean of 18.61 days and standard deviation 13.62 days.

To compute the discretised delay vectors from infection to case confirmation, and for infection to reported death, we use Monte Carlo integration to discretise and sum the incubation period with the relevant delay.

*Generation interval.* We take an estimate for the generation interval from a meta-analysis (*14*). We use Monte Carlo integration to discretise this delay.

Table 1 lists the delay distributions that we use, as well as their sources.

**Table 1: Table of epidemiological parameters, their distributional forms and their source.**

| Delay | Distributional form of delay | Source |
|---|---|---|
| Generation interval | Gamma(mean=4.83, sd = 1.73) | Meta-analysis (*14*) |
| Incubation period | Gamma(mean=5.53, sd = 4.73) | Meta-analysis (*14*) |
| Onset to reported death | Gamma(mean=18.61, sd = 13.62) | Linelist (Sec. 1.2) |
| Onset to case confirmation | Gamma(mean=5.28, sd = 3.75) | Linelist (Sec. 1.2) |

### Supplementary text

### 1. Validation

#### 1.1. Sensitivity analysis

Sensitivity analysis reveals the extent to which results depend on uncertain parameters and modelling choices, and can diagnose model misspecification and excessive collinearity in the data (*16*). We vary many of the components of our model and recompute the NPI effectiveness estimates[a]. Overall, we perform 17 sensitivity analyses with 86 experimental conditions. Table S1 summarises our sensitivity analyses and their categories (matching Fig. **3**).

**Table S1: Sensitivity analysis categories.**

| Category Name | Analyses |
|---|---|
| Delay distributions | Generation interval mean (Figure S1) |
| | Infection to confirmation delay (Figure S2) |
| | Infection to reported-death delay (Figure S3) |
| Data | Bootstrap (Figures S4,S5, and S6) |
| | Maximum VOC permitted (Figure S7) |
| | Schools delayed (Figure S8) |
| Prior distributions | Infection noise scale (Figure S9) |
| | $\sigma_R$ prior (Figure S10) |
| | Output noise scale prior (Figure S11) |
| | Intervention prior (Figure S12) |
| | $\tilde{R}_{0,l}$ mean (Figure S13) |
| | $\tilde{R}_{0,l}$ scale (Figure S14) |
| | Seeded infections prior (Figure S15) |
| Model structure | Random walk period (Figure S16) |
| | Seeding days (Figure S17) |
| Unobserved factors | NPI leaveout (Figures S18,S19,S20, and S21) |
| | Time-varying IFR/IAR adjustment (Figure S22) |

[a] As in the main text, NPI effectiveness is displayed as the percentage reduction in $R_t$, computed as $1-\exp(-\beta_i)$.

###### 1.1.1. *Sensitivity to delay distributions*

Figures S1, S2 and S3 show the sensitivity of the NPI effectiveness estimates to the means of the distributions of the generation interval, the delay between infection and case reporting, and the delay between infection and death reporting.

Consistent with Flaxman et al. (*10*), Brauner et al. (*1*) and Sharma et al. (*17*), we find that a shorter mean generation interval implies smaller values of $R_t$ when the epidemic is growing, and therefore lower effectiveness estimates. Furthermore, the sensitivity to the delay between infection and case reporting appears to be larger than the sensitivity to the delay between infection and death reporting.

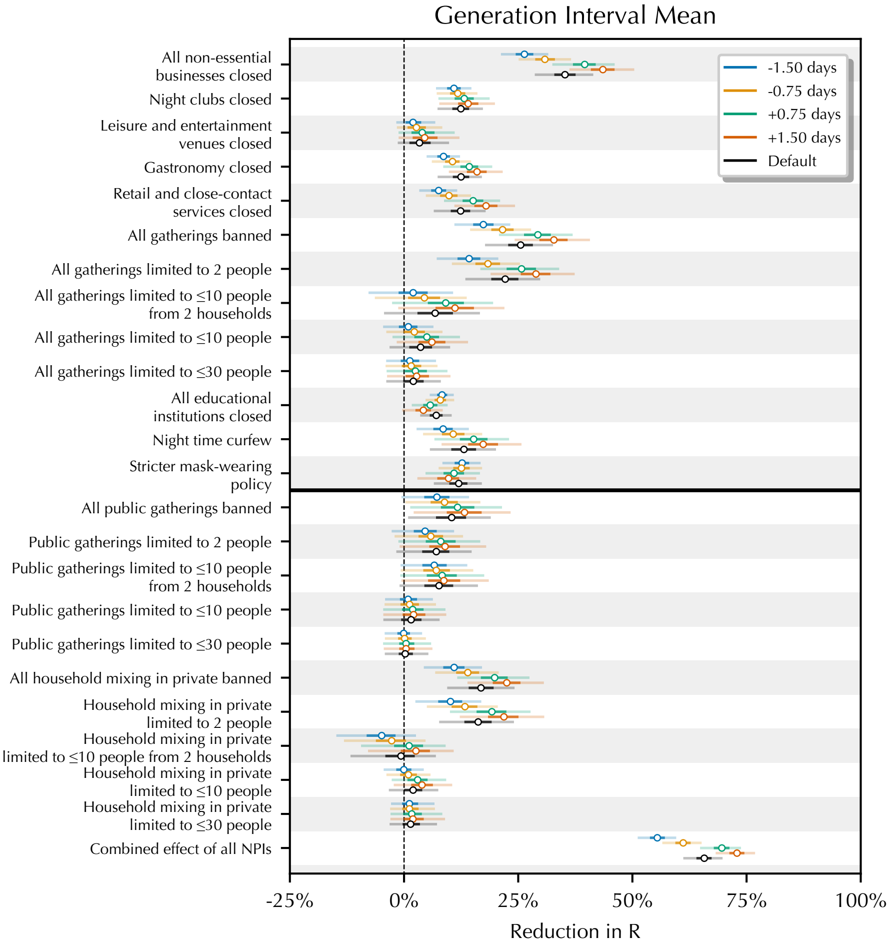

**Figure S1: Sensitivity of NPI effect estimates to the mean of the generation interval.**

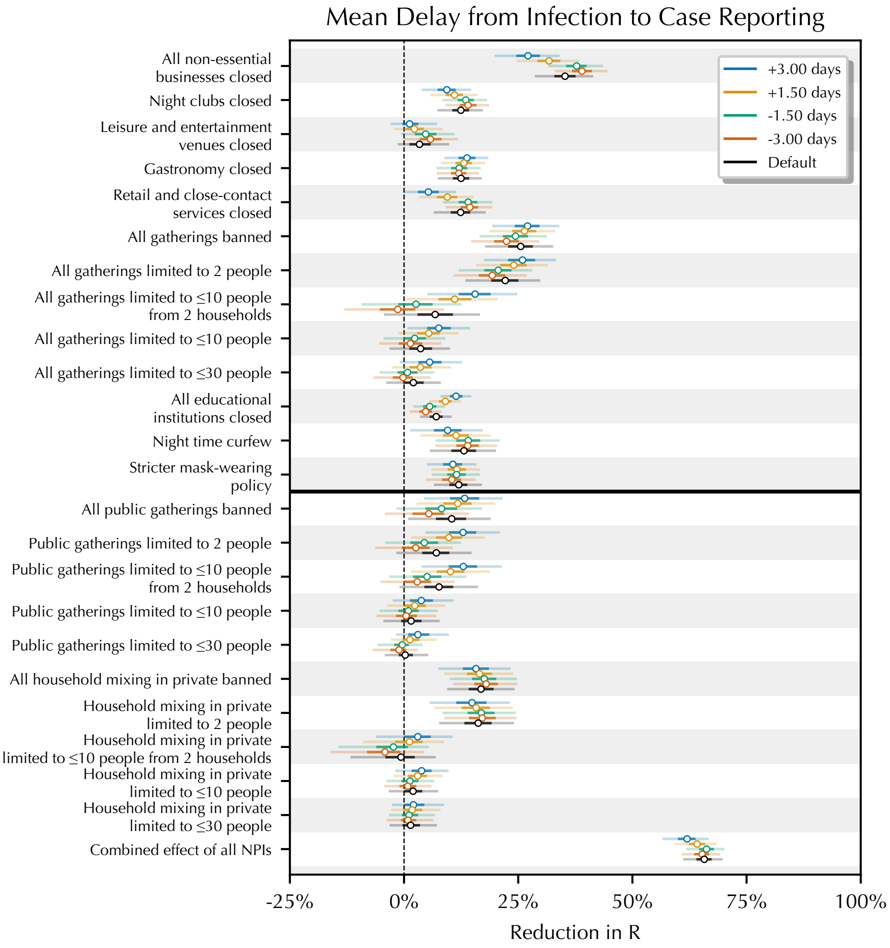

**Figure S2: Sensitivity of NPI effect estimates to the mean of the delay from infection to case reporting.**

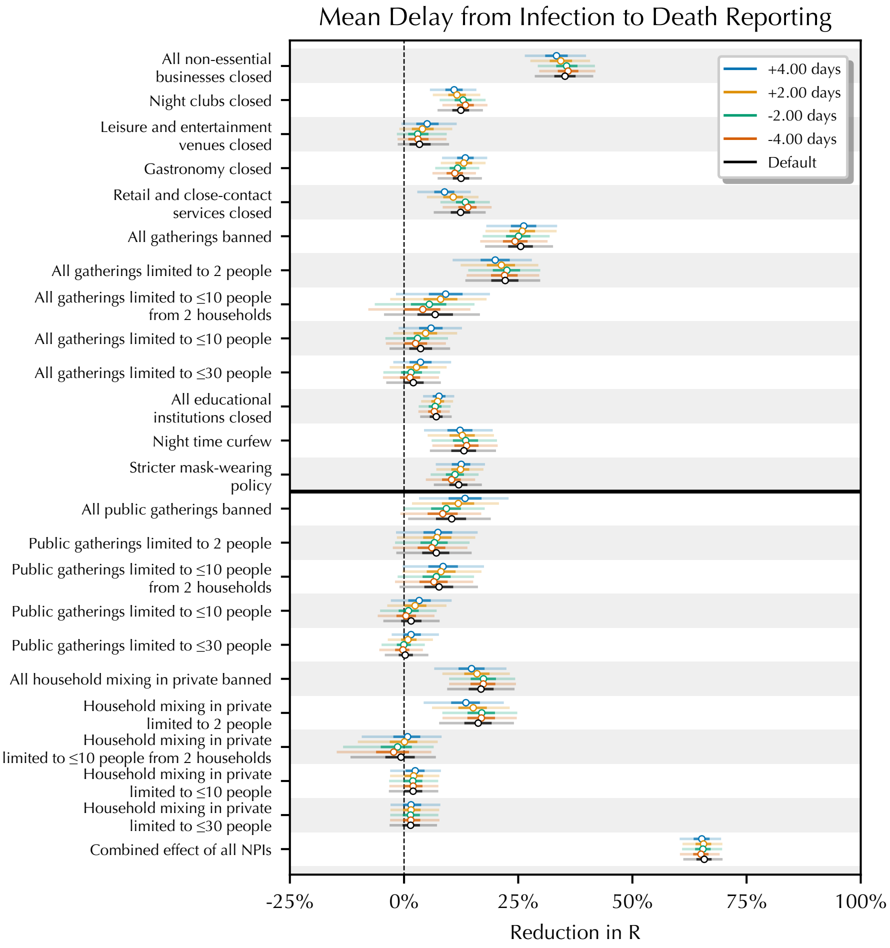

**Figure S3: Sensitivity of NPI effect estimates to the mean of the delay from infection to death reporting.**

*1.1.2. Sensitivity to data*

Figures S4, S5 and S6 show the sensitivity of NPI effect estimates when bootsrapping over the locations in our dataset. For each random seed, we sample 114 locations **with replacement** from our pool of locations. Bootsrapping assesses the extent to which our NPI effect estimates depend on the specific locations that were included. Each bootstrap typically samples around 75 *unique* locations.

Figure S7 shows the sensitivity of NPI effect estimates when the maximum permitted fraction of cases attributed to the b.1.1.7 Variant of Concern (VOC) is changed. This sensitivity analysis only affects locations within England.

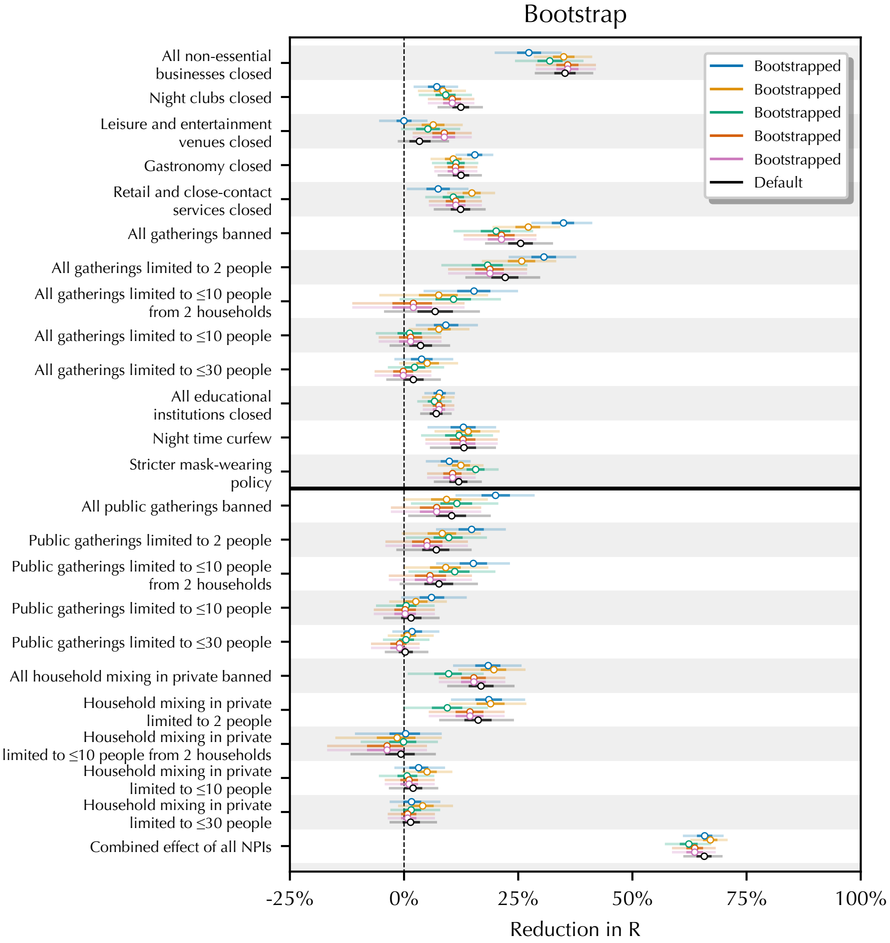

**Figure S4: Sensitivity of NPI effect estimates using random bootstrapped sets of regions.**

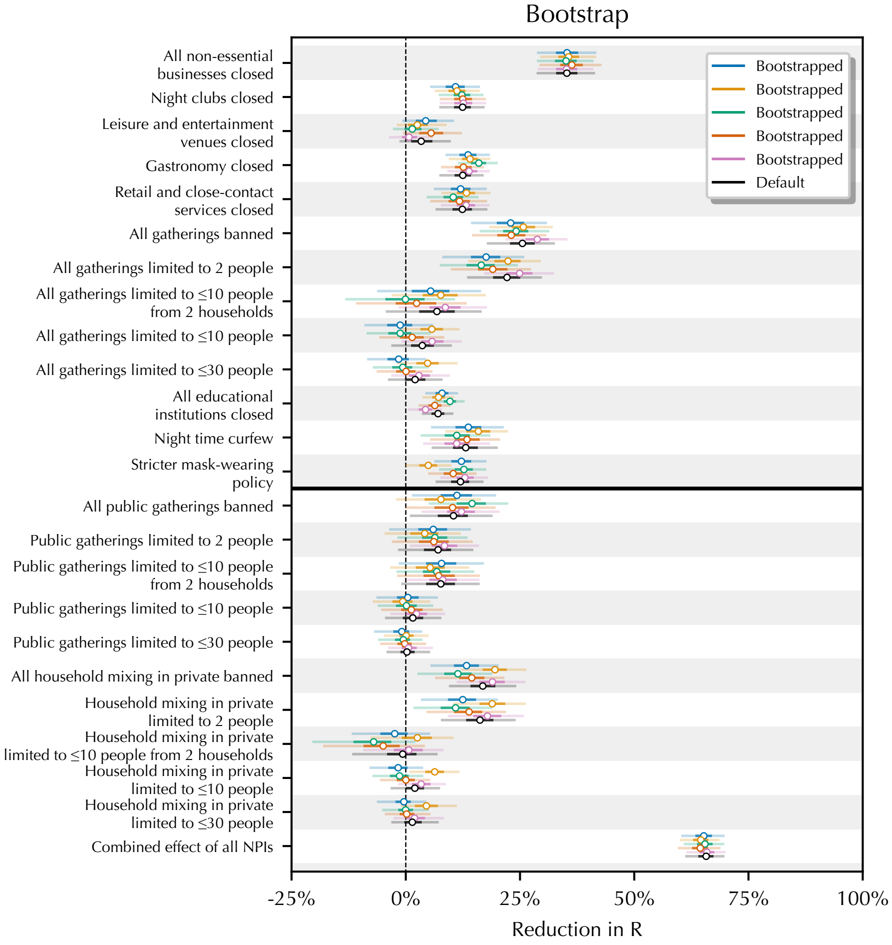

**Figure S5: Sensitivity of NPI effect estimates using random bootstrapped sets of regions.**

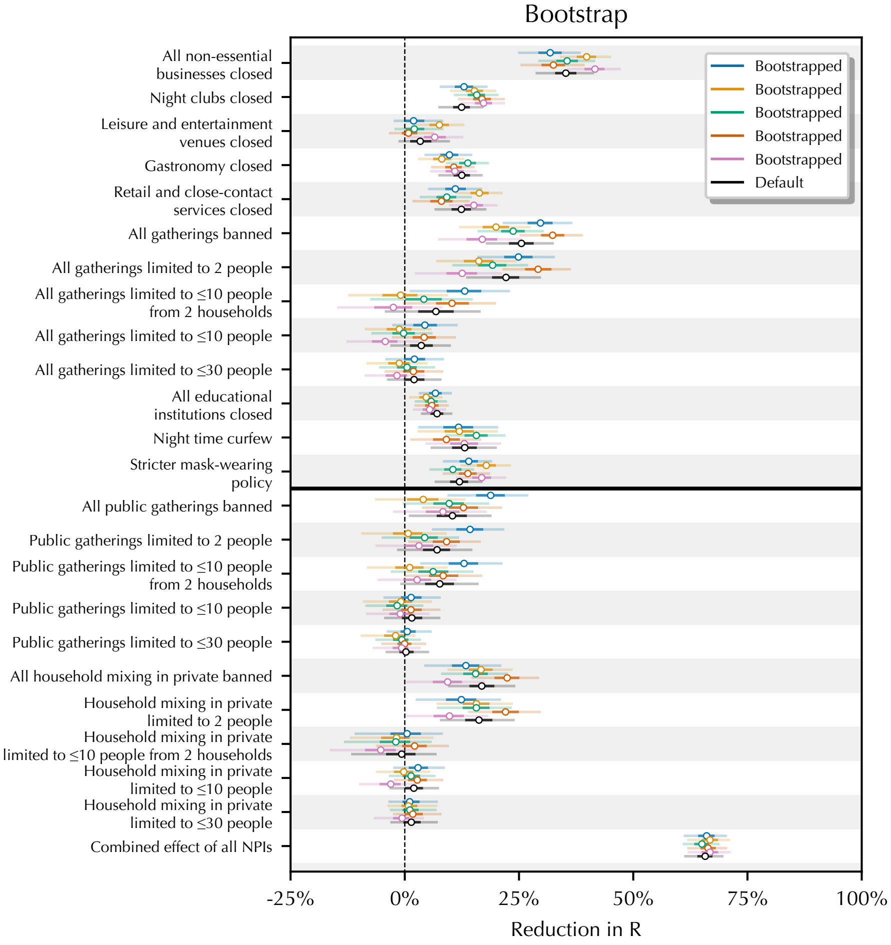

**Figure S6: Sensitivity of NPI effect estimates using random bootstrapped sets of regions.**

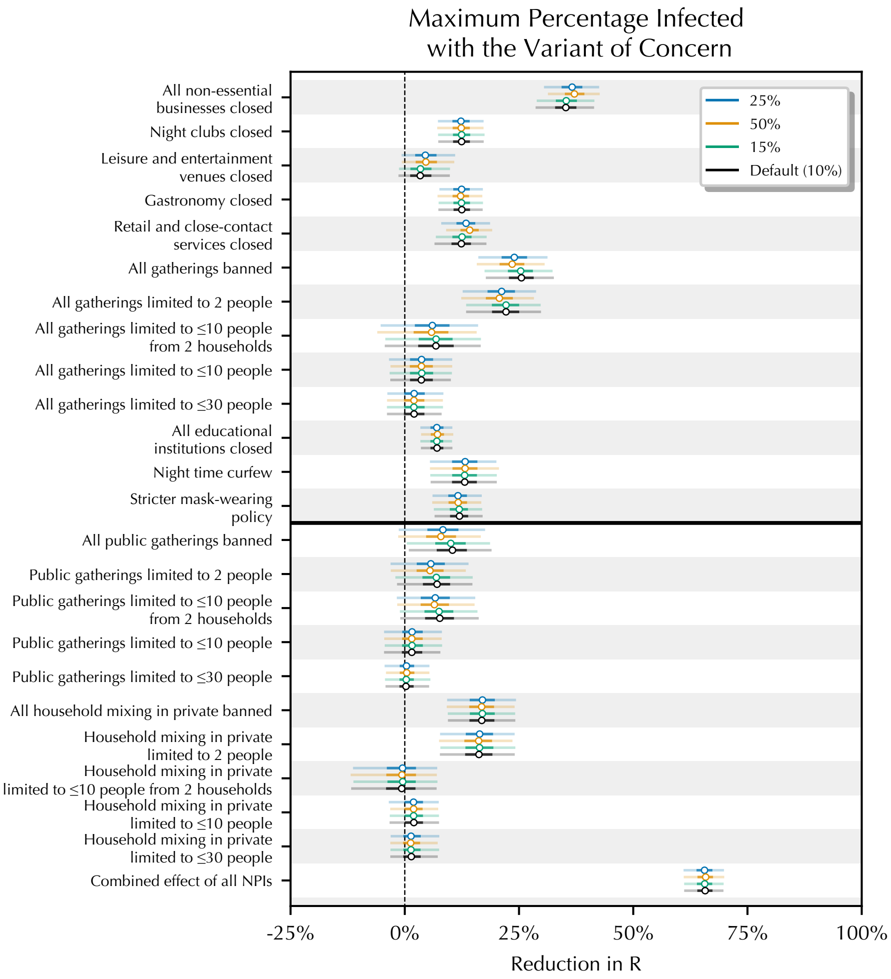

**Figure S7: Sensitivity of NPI effect estimates to changing the maximum proportion of the variant of concern allowed before the window of analysis is ended. Note: this sensitivity analysis only affects England.**

*1.1.3. The individual effects of school and university closures are sensitive*

Some evidence points towards underreporting of COVID-19 cases amongst school-age children. For example, children may be more likely to be asymptomatic, and thus more likely to remain untested (*18*). An important consequence of this is that the effect of school openings and closures may be lagged. For example, suppose COVID-19 infections amongst school-age children remain undetected, and that school opening increases transmission. Then, if schools were to open, the number of reported cases in a location would only rise once children, who were infected when schools opened, subsequently infect members of the adult population. To investigate this, we delay the intervention time series for school closure NPIs by approximately one generation interval–the average time between successive infections in a transmission chain.

Figure S8 shows the sensitivity of the school closure and universities closed NPIs when we delayed the school NPI time series by one mean generation interval. While the total effect of closing *all* education institutions remains comparatively stable, the relative effectiveness of school and university closures shifts clearly. If we believed that delay between school closures and reported cases and deaths is similar to other NPIs, we would conclude that transmission in universities is more significant than in schools. However, if we believed that school closures had an *additionally* delayed effect on reported cases and deaths in comparison to other NPIs, we would conclude the opposite. Instead, we conclude that we cannot robustly disentangle the *individual* effects of school closures and university closures; as such, we report their *combined* effect, which is more stable.

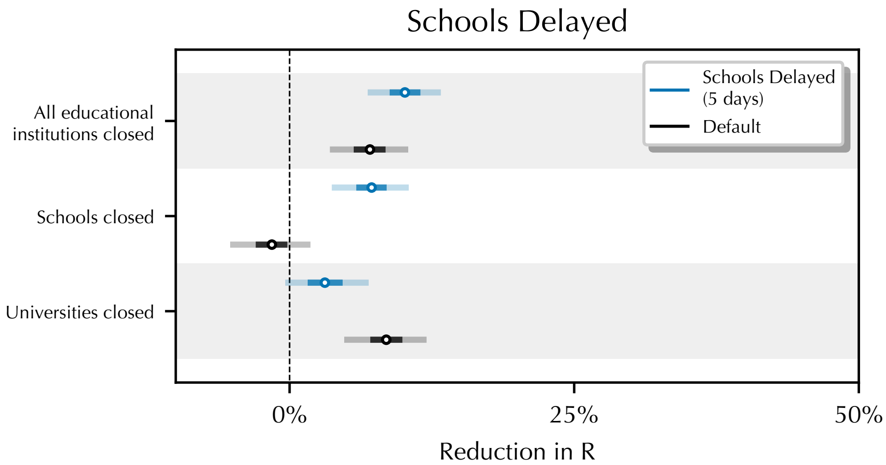

**Figure S8: Sensitivity of effect estimates for education NPIs when delaying school openings and closures by one generation interval (5 days).**

*1.1.4. Sensitivity to prior distributions*

Figure S9 shows the sensitivity of NPI effect estimates to the infection noise scale.

Figure S10 shows the sensitivity of NPI effect estimates to the prior over the random walk noise scale, $\sigma_R$.

Figure S11 shows the sensitivity of NPI effect estimates to the prior over the country-specific output noise scales, $\Psi_c^{(C)}$ and $\Psi_c^{(D)}$.

Figure S12 shows the sensitivity of NPI effect estimates to the prior over the NPI effects.

Figure S13 shows the sensitivity of NPI effect estimates to the mean of the prior over $\tilde{R}_{0,l}$. Recall that $\tilde{R}_{0,l}$ is the reproduction number at the start of the window of analysis, supposing no NPIs are active.

Figure S14 shows the sensitivity of NPI effect estimates to the scale/standard deviation of the prior over $\tilde{R}_{0,l}$. Recall that $\tilde{R}_{0,l}$ is the reproduction number at the start of the window of analysis, supposing no NPIs are active.

Figure S15 shows the sensitivity of NPI effect estimates to the scale of the prior over the seeded infections. All priors are of the form $\text{Lognormal}(0, \sigma)$.

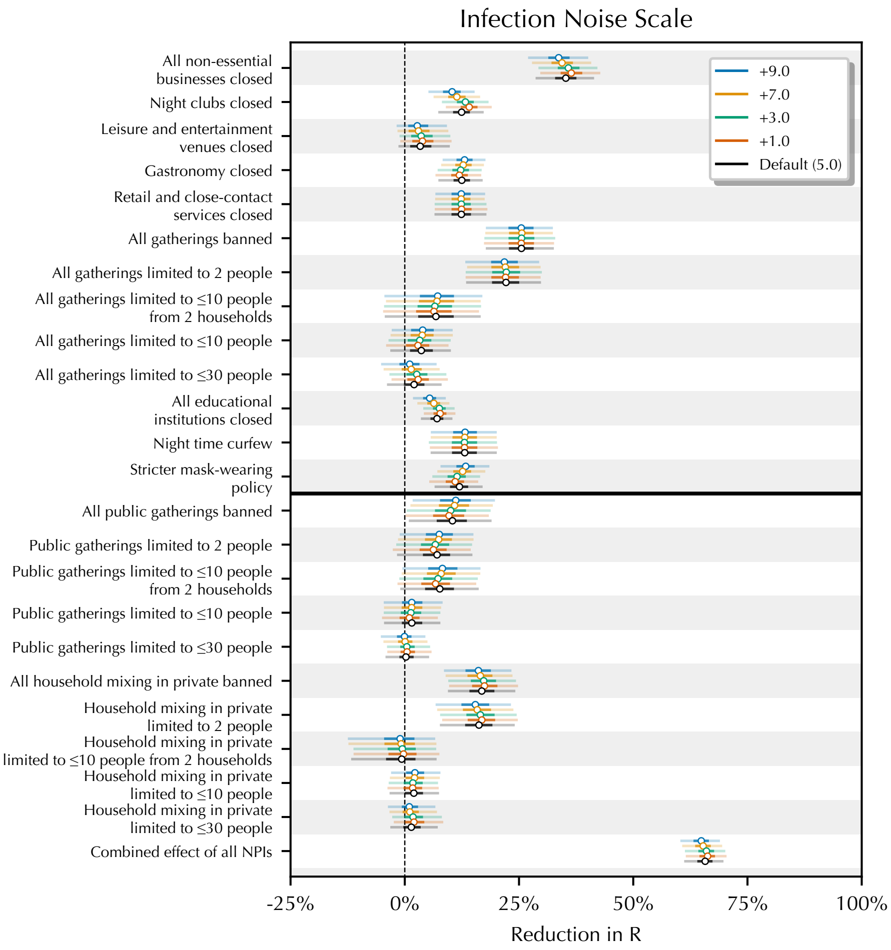

**Figure S9: Sensitivity of NPI effect estimates to the infection noise scale.**

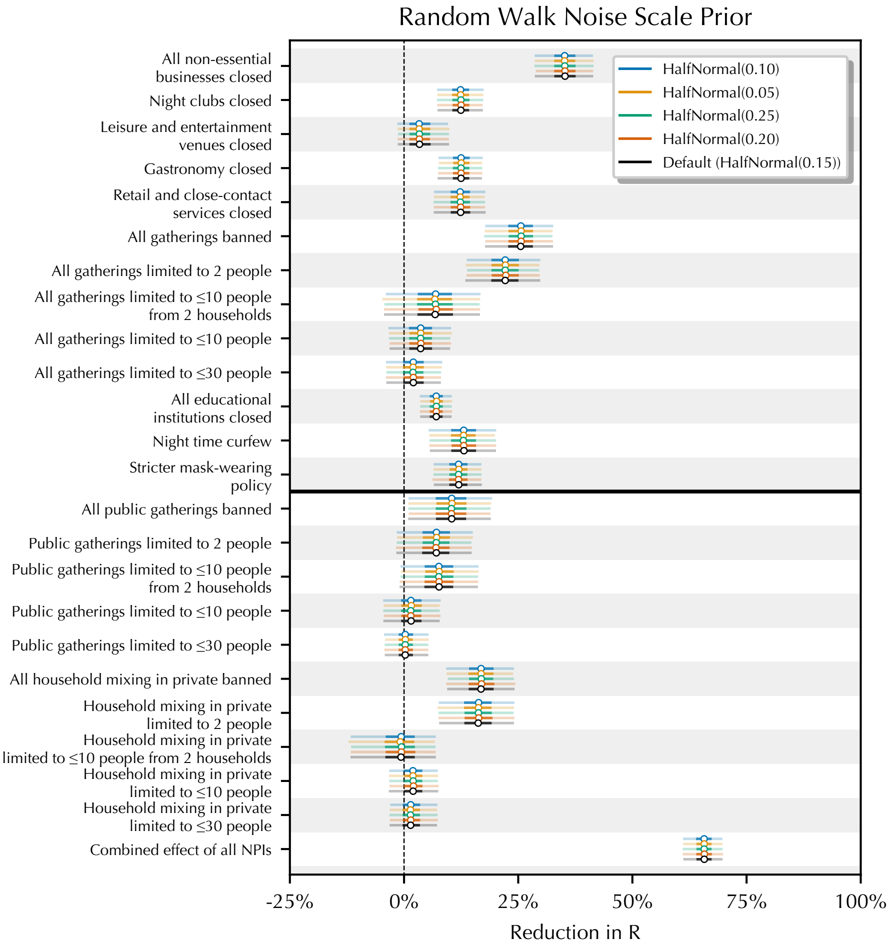

**Figure S10: Sensitivity of NPI effect estimates to the prior over the scale of the latent random walk.**

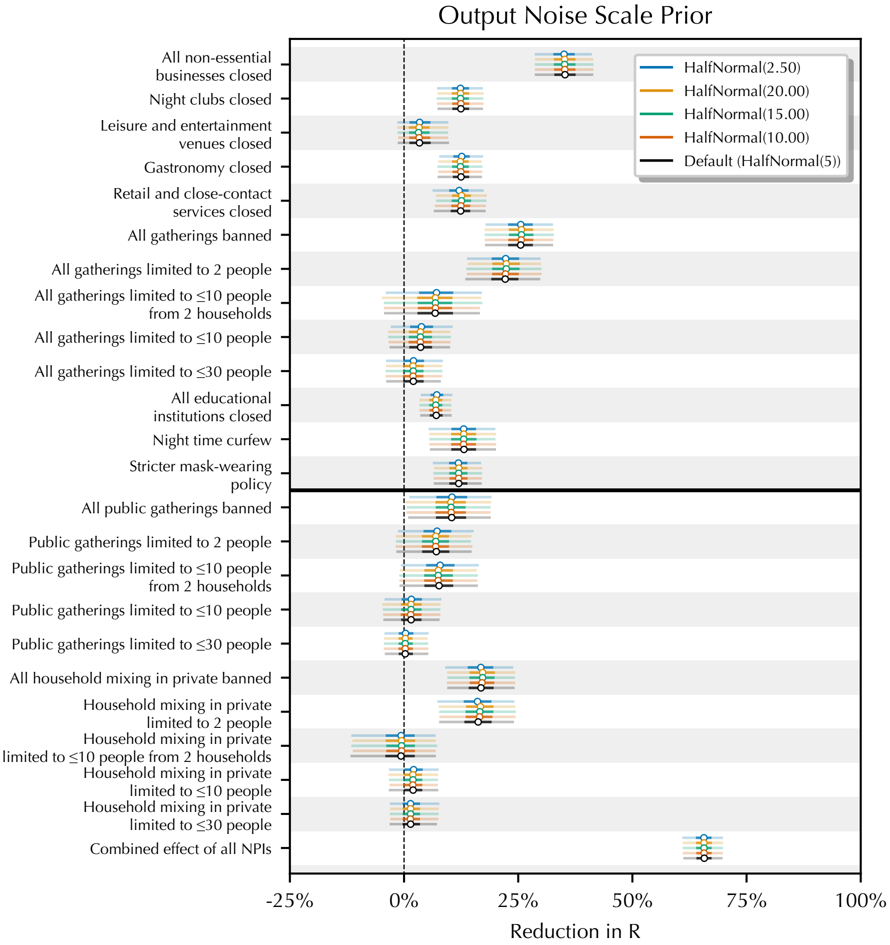

**Figure S11: Sensitivity of NPI effect estimates to the prior over the output noise scale.**

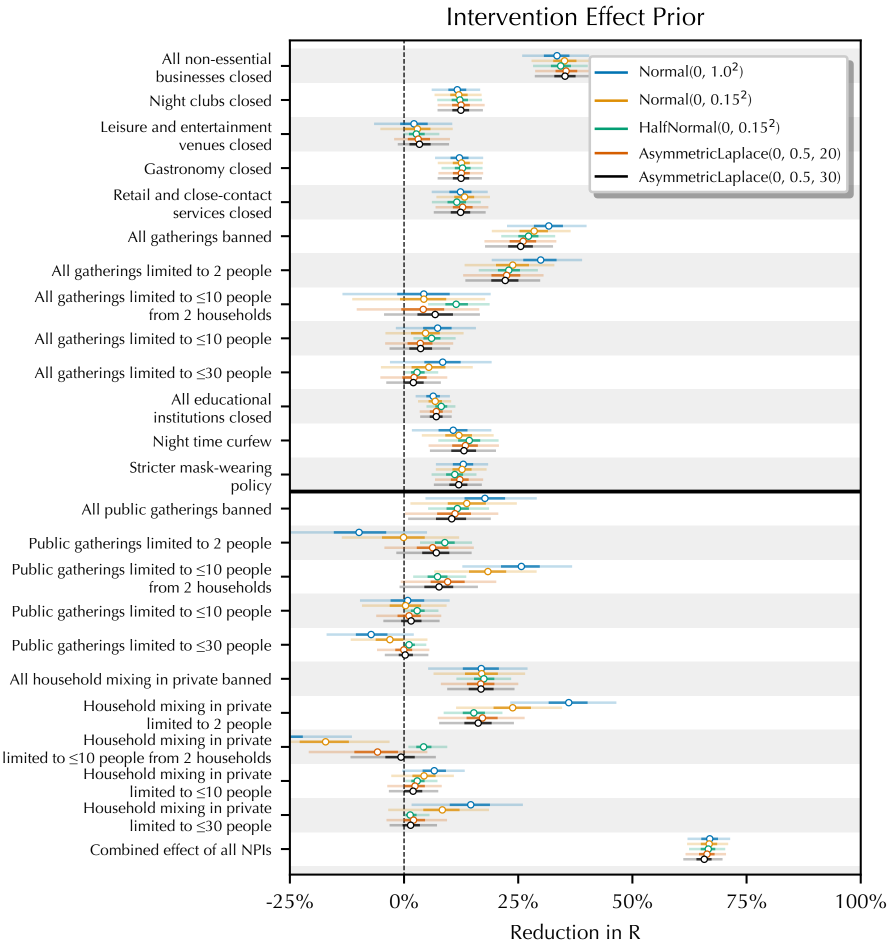

**Figure S12: Sensitivity of NPI effect estimates to the prior over the intervention effect.**

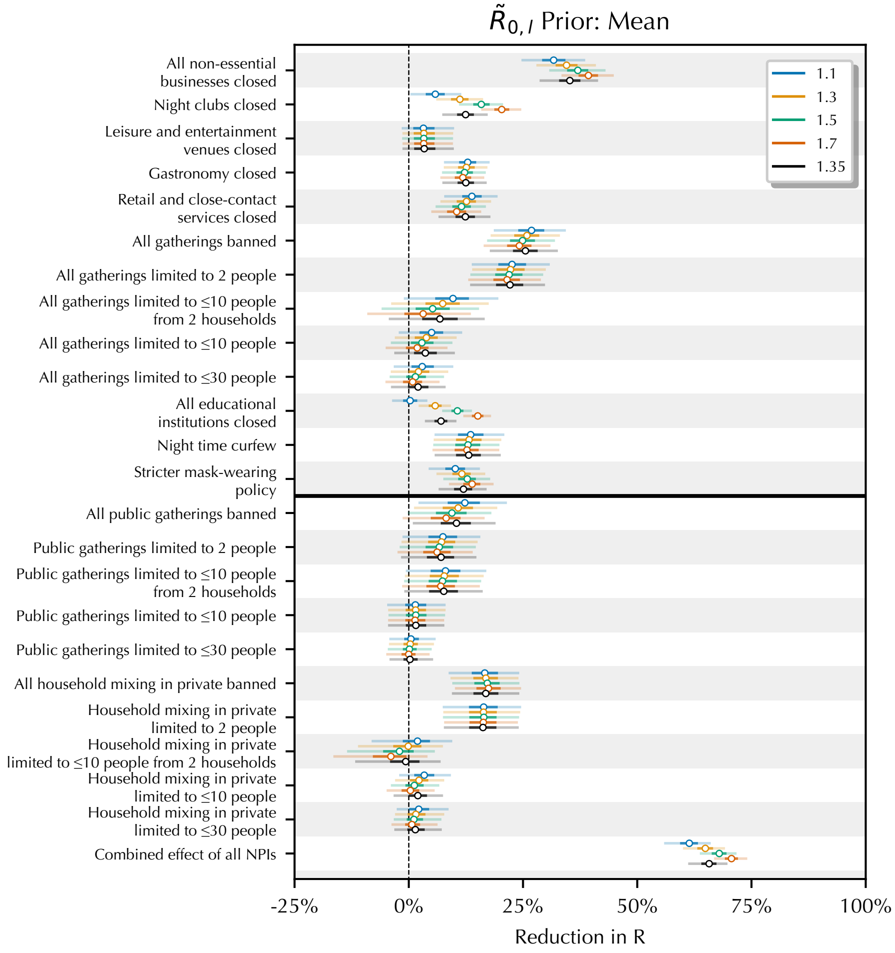

**Figure S13: Sensitivity of NPI effect estimates to the mean value of the prior over $\tilde{R}_{0,l}$ i.e., the value of the reproduction number at the start of the window of analysis, supposing no NPIs were active.**

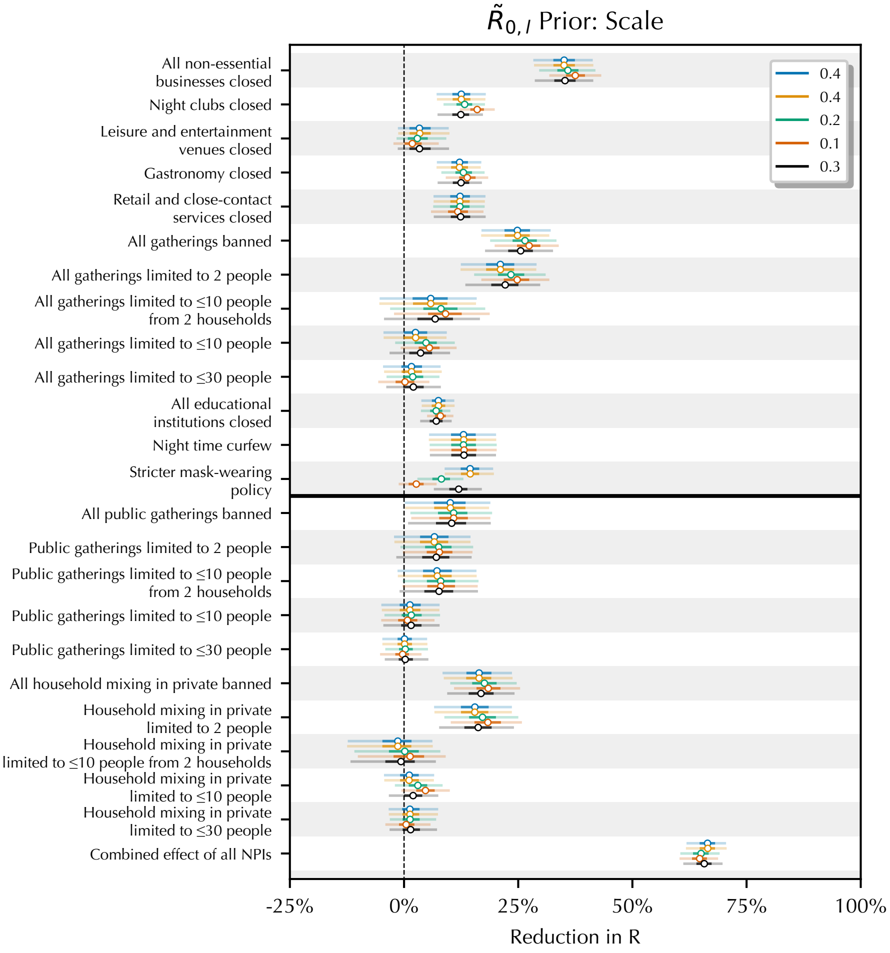

**Figure S14: Sensitivity of NPI effect estimates when the variability/scale of the prior over $\tilde{R}_{0,l}$ i.e., the value of the reproduction number at the start of the window of analysis, supposing no NPIs were active.**

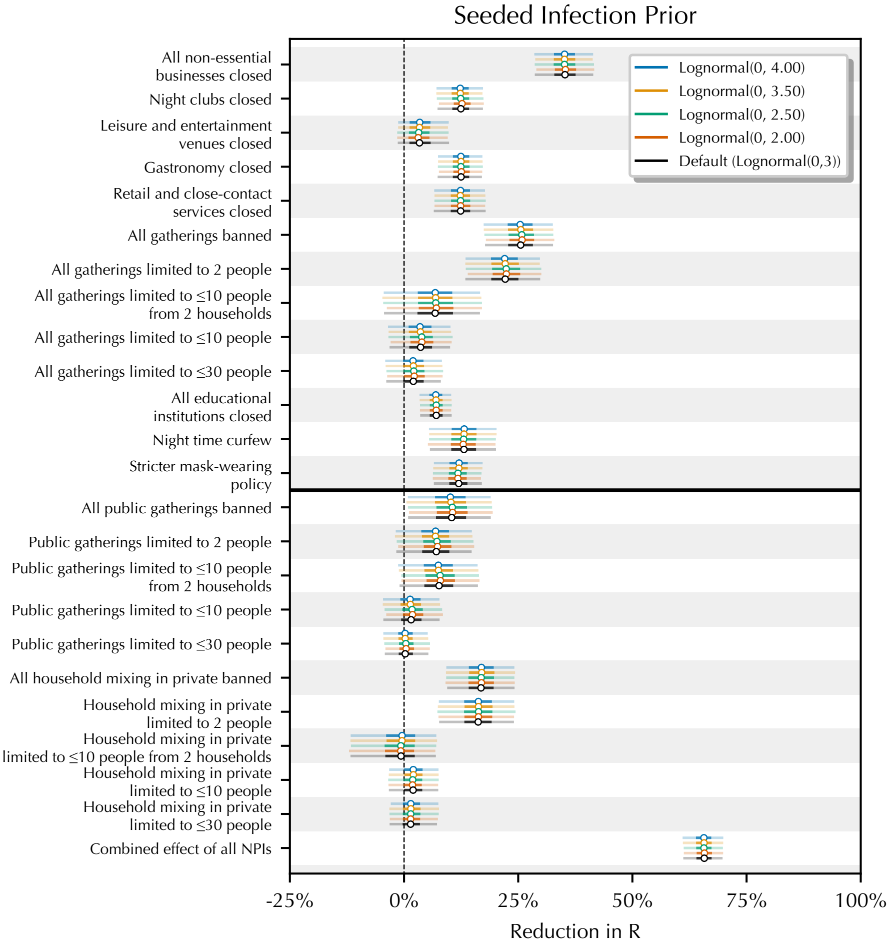

**Figure S15: Sensitivity of NPI effect estimates to the prior over over the seeded infections.**

*1.1.5. Sensitivity to model structure*

Figure S16 shows the sensitivity of NPI effect estimates to the period of the latent random walk. If the period is 5 days, for example, then the value of $R_{t,l}$ may change (without a change of interventions) every 5 days.

Figure S17 shows the sensitivity of NPI effect estimates to the number of days of seeded infections.

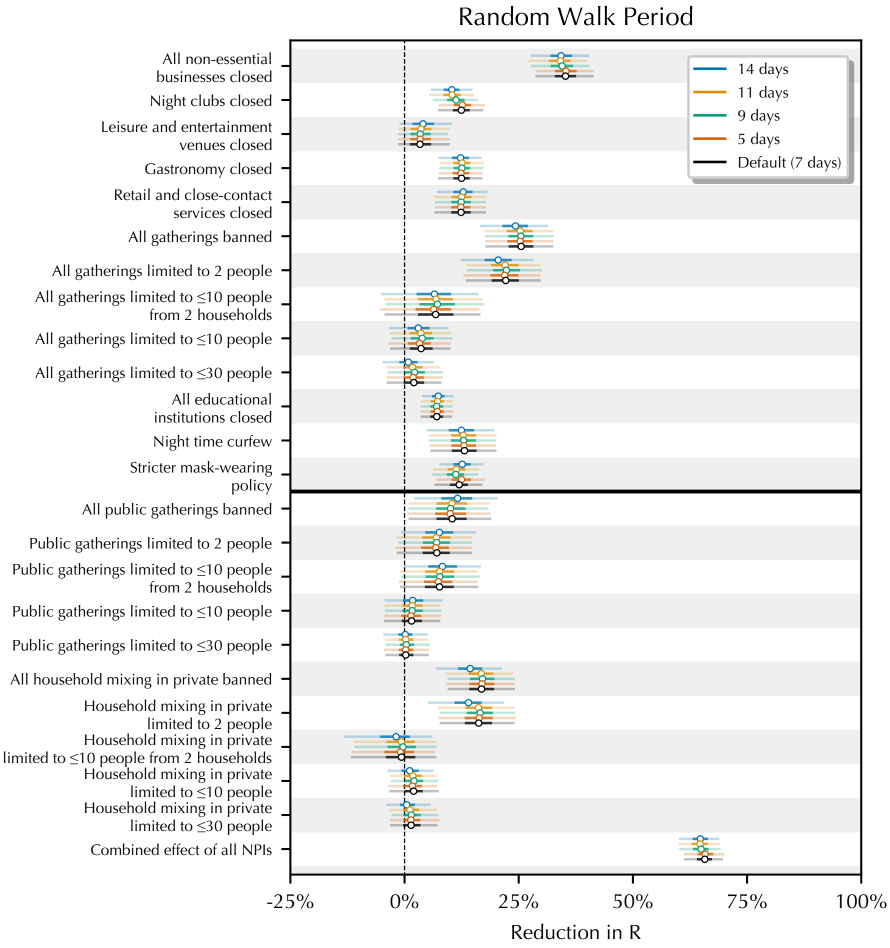

**Figure S16: Sensitivity of NPI effect estimates to the period of the random walk.**

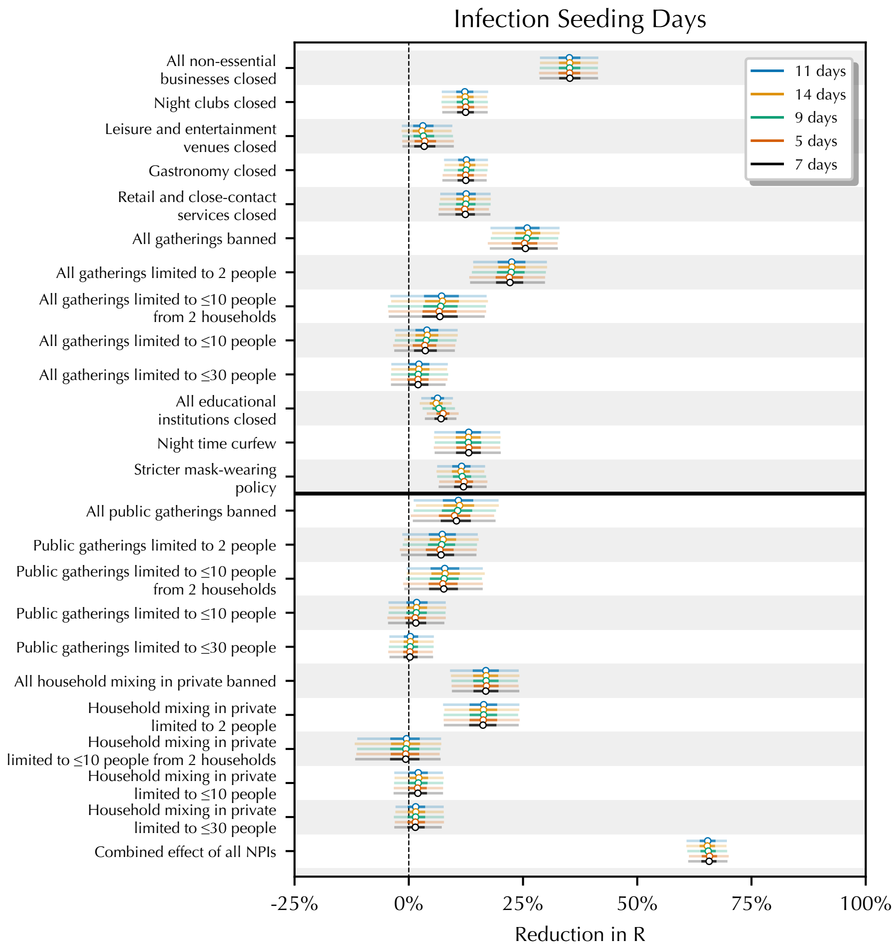

**Figure S17: Sensitivity of NPI effect estimates to the number of days of seeded infections.**

*1.1.6. Sensitivity to unobserved factors*

Our data neither captures all of the NPIs that were implemented nor directly measures voluntary behaviour changes. Since these factors influence $R_t$, we must be wary of their effect being attributed to the observed NPIs. Unobserved factors can bias results if their timing is correlated with the timing of the observed NPIs (*19*).

We investigate this by assessing how much effectiveness estimates change when previously when observed factors are excluded. This is best practice for addressing unobserved factors, such as confounders (*20, 21*), and follows Sharma et al. (*17*). Note: if we remove a component of a hierarchical NPI, we do not show the same hierarchical NPI, since its value may no longer be defined. For example, if we were to exclude retail closures, the effect of closing all non-essential businesses is not displayed.

Figures S18, S19, S20, and S21 shows NPI effectiveness estimates when previously observed NPIs are excluded in turn. Considering that some of the excluded NPIs have strong estimated effects when included, and are correlated with other NPIs, this degree of robustness is encouragingly high.

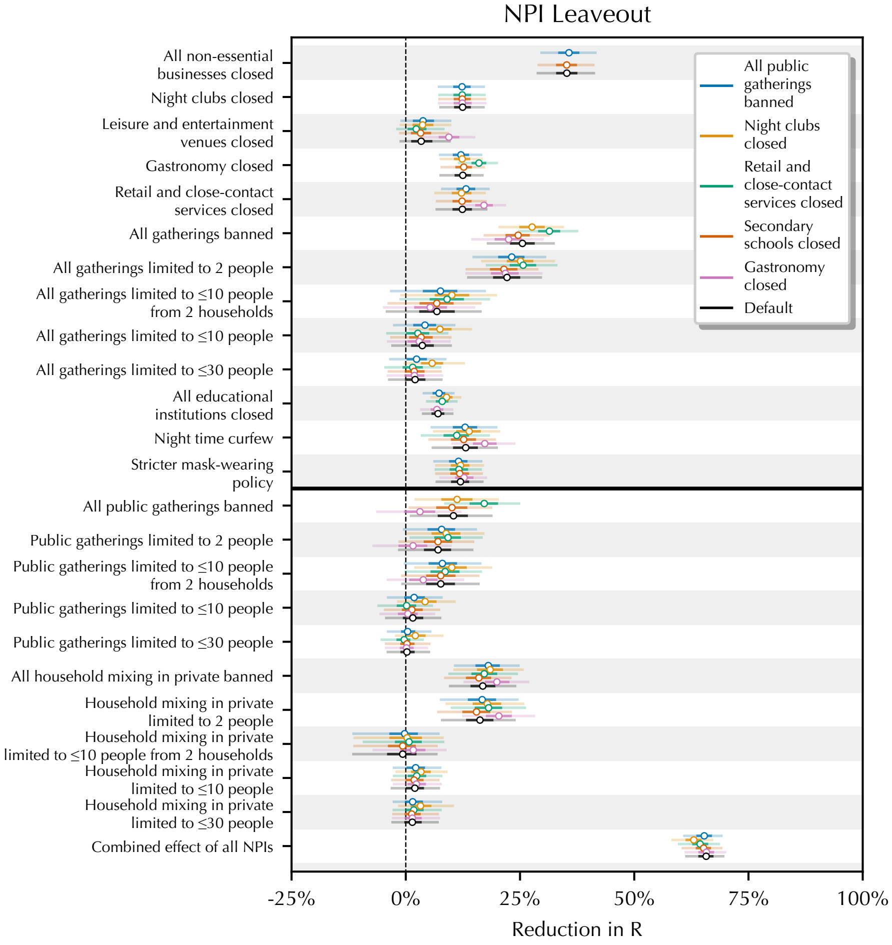

**Figure S18: Sensitivity of NPI effect estimates to leaving out recorded interventions.**

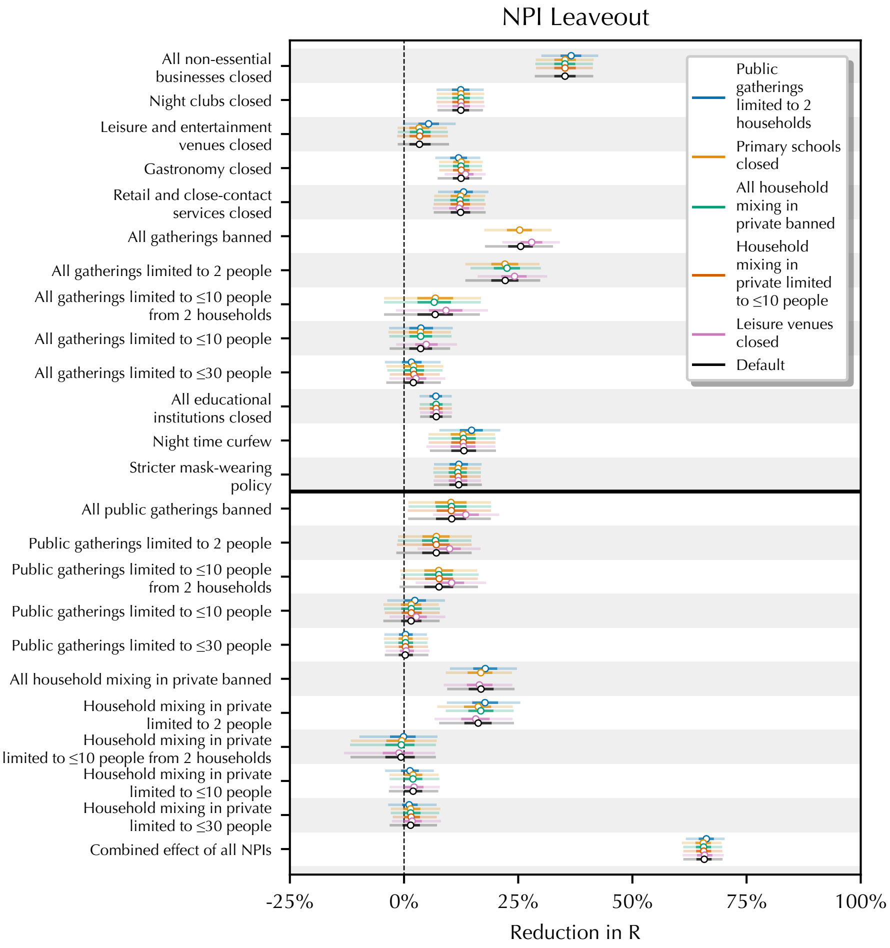

**Figure S19: Sensitivity of NPI effect estimates to leaving out recorded interventions.**

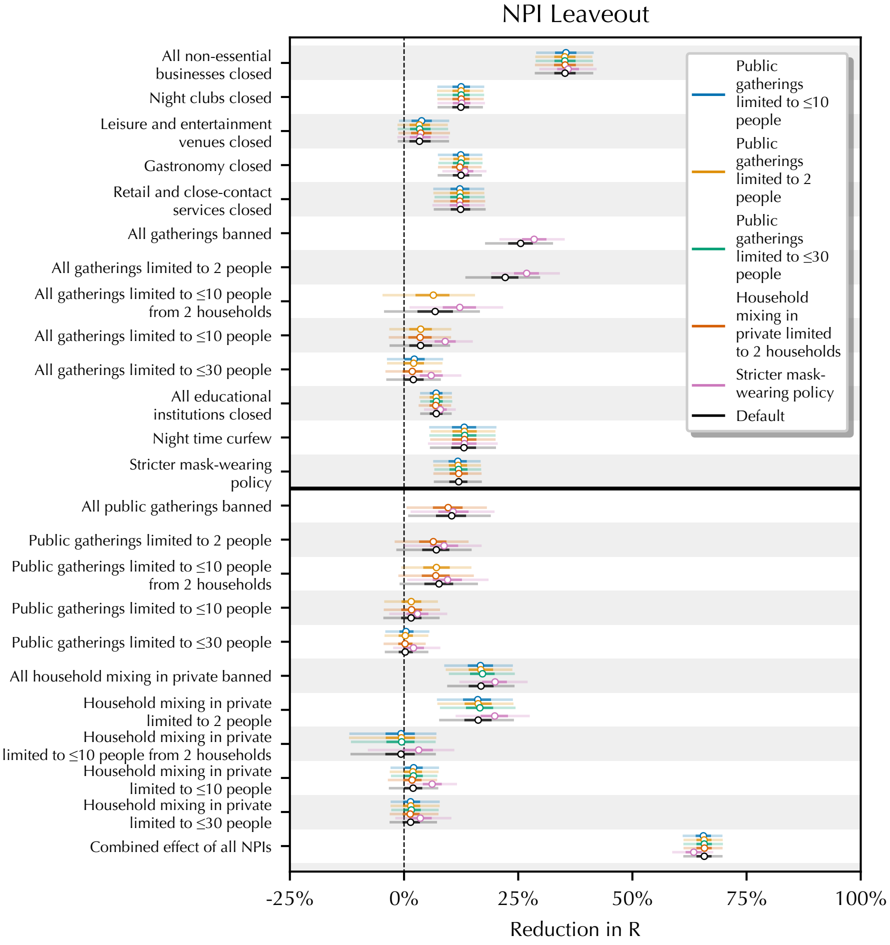

**Figure S20: Sensitivity of NPI effect estimates to leaving out recorded interventions.**

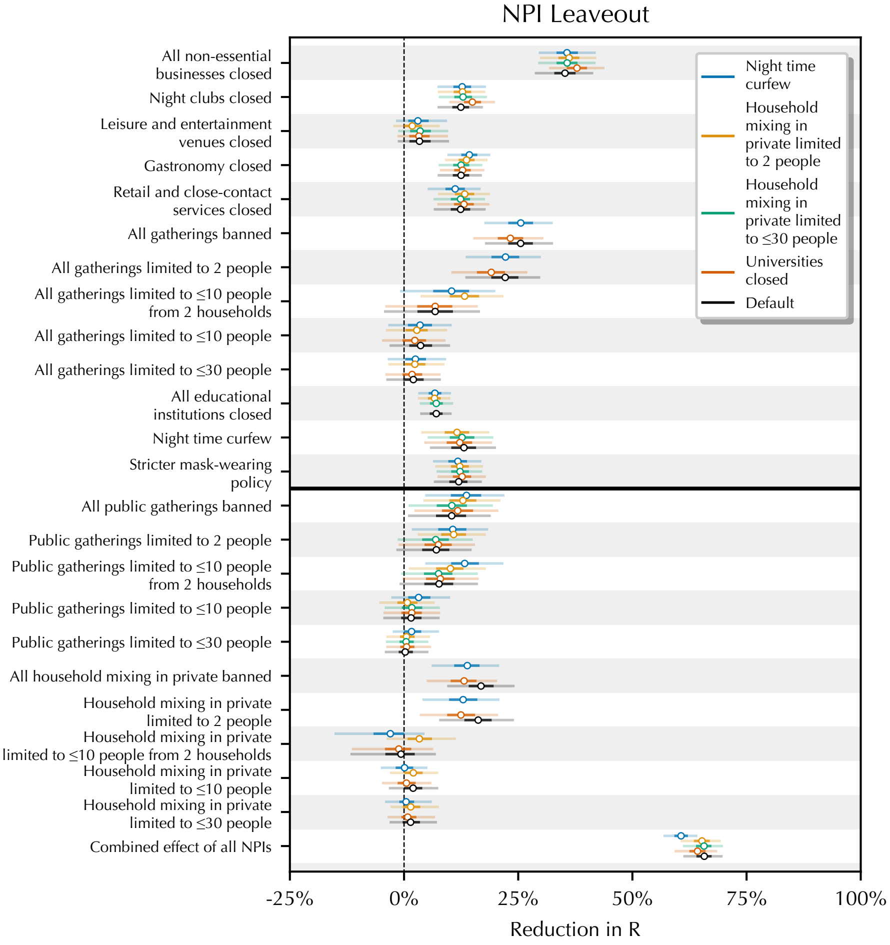

**Figure S21: Sensitivity of NPI effect estimates to leaving out recorded interventions.**

##### 1.1.7. *Sensitivity to Changing IFR and IAR*

Recall that our model assumes a time-constant Infection-Fatality Rate (IFR)–the proportion of infections that result in fatalities–and a time-constant Infection-Ascertainment Rate (IAR)–the proportion of infections that are confirmed as COVID-19 cases. In reality, the IFR and IAR may change over time. For example, improvements in COVID-19 treatment and increasing testing capacity may cause the IFR to decrease and the IAR to increase. Therefore, we assess the sensitivity of our NPI effectiveness estimates when accounting for the time-varying IFR and IAR.

In particular, we use estimates of the IFR and the IAR in England, taken from Mishra et al. (*7*). The time-varying estimates of IFR/IAR are estimated using seroprevalence data from ONS (*8*) and REACT (*9*) along with case and death time series for England. While the trends in the IAR and IFR are unique to every country, the trend in England is indicative of how, and how much the IFR and IAR may change in other countries. Thus, to investigate the time-constant IFR and IAR assumption, we apply the estimates from England to *all* of our locations. Specifically, instead of Eq. (8), we use:

$$N_{t,l}^{(C)} = \widehat{\text{IAR}}_{t,\text{england}} \cdot N_{t,l}, \quad \text{and } N_{t,l}^{(D)} = \widehat{\text{IFR}}_{t,\text{england}} \cdot N_{t,l}. \tag{A.1}$$

$\widehat{\text{IAR}}_{t,\text{england}}$ is the time-varying estimate for the ascertainment rate in England, while $\widehat{\text{IFR}}_{t,\text{england}}$ is the corresponding infection fatality rate estimate.

Figure S22 shows the NPI effectiveness estimates when adjusting the time-varying IFR/IAR. We find low sensitivity to this change, suggesting that the time-constant IFR and IAR is not a key limitation of this work.

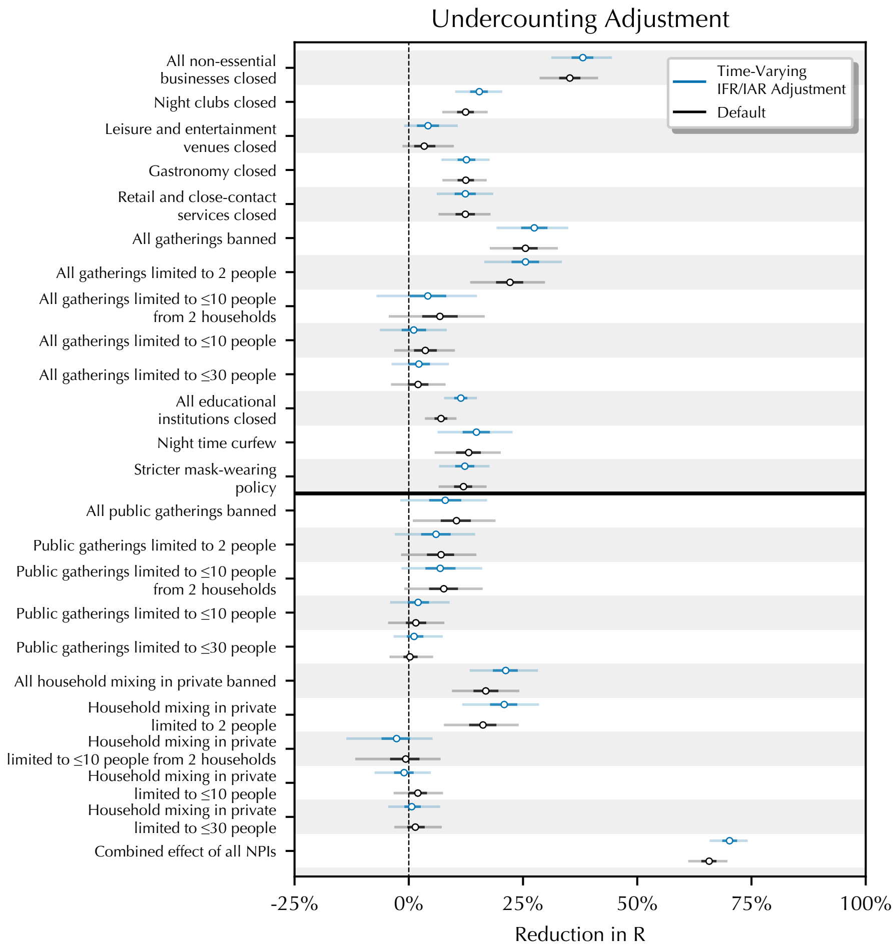

**Figure S22: Sensitivity of NPI effect estimates when adjusting for time-varying case undercounting (i.e., a time-varying ascertainment rate), as well as a time-varying infection-fatality rate.**

#### 1.2. Simulation study

We now assess whether our model can correctly recover the ground truth NPI effects. We use our model to simulate synthetic epidemics for all 114 regions, keeping the original timing and ordering of interventions but assigning hypothetical effect sizes to each intervention. If we are able to recover the hypothetical effect sizes, this suggests that we are able to recover the "ground truth" NPI effectiveness estimates. We assigned small effect sizes to all but one NPI, to which we assigned a (median) 29% reduction in $R_t$. Note that we use the real intervention dates to ensure that our simulation is reflective of reality, and the simulated NPIs have the same collinearity as the real NPIs.

Furthermore, in reality, we cannot record all interventions present in a local area. To reflect this in our simulation, and assess whether our NPI estimates have been biased by the omitted variable, we include an additional location-specific NPI. This NPI is introduced at a random time in each region and has a different effect in every region. **This NPI is not observed by our model**. In the simulation, this NPI's effect is drawn from a diffuse distribution bounded between 0% and 100%, with mean 27%. In reality, omitted variables may include, for instance, spontaneous behaviour change in response to government messaging and could bias the effect size estimates of our recorded NPIs.

Next, we fit our model to these synthetic datasets, using 50 different simulations for each setting. As shown in Figure S23, the estimates from the fitted model are close to the simulated effect size for the dominant NPI, and for all others, the effect size is small with overlapping intervals. This analysis provides evidence that our model can recover "ground truth" NPI effectiveness estimates, even in the presence of unrecorded interventions.

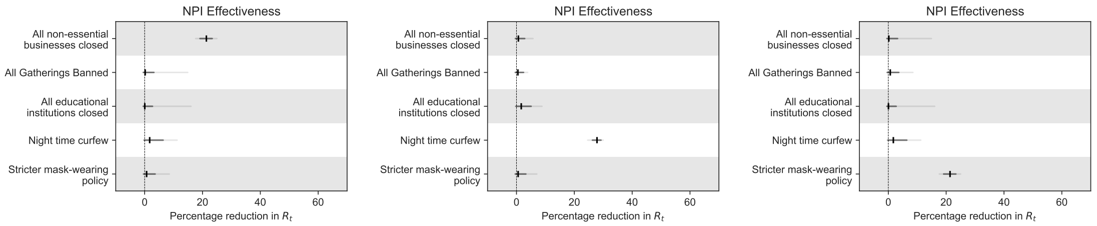

**Figure S23: Estimated effect sizes from simulated data. Right to left, three separate simulation settings. For each setting, one NPI is assigned a large effect (median 29%). Also, for each setting, we perform 50 model runs, drawing 50 datasets from the simulation distribution and running our model on each of them. Each panel shows effect sizes from the main model fits across the 50 simulations. We show the median point estimate and 95% credible interval taken across the 50 runs.**

#### 1.3. Prior predictive checks

Graphical prior predictive checks help us to understand whether our data generation process produces data that obeys physical constraints and matches our intuition (*22*). Since we have included a latent random walk, it is important to understand the prior predictive distribution on $R_t$.

Figure S24 shows prior predictive distributions for $R_t$, produced using our NPI intervention timing dataset. Note that the prior on $R_{t,l}$ depends on the active interventions. Under the prior, the uncertainty in $R_{t,l}$ increases over time as noise in the latent random walk accumulates. However, since we assume that the latent random walk is equally likely to increase and decrease transmission, the *median* value of $R_{t,l}$ changes only when the active interventions change in each particular location. Since we have assumed that interventions are more likely to decrease transmission than increase transmission, under the prior distribution, we find that $R_{t,l}$ decreases when NPIs are implemented–see e.g., Abruzzo at the end of October.

These prior checks show the appropriateness of our model specification with regards to prior assumptions. The simulated values are likely to lead to a second wave. Following interventions from October, $R_{t,l}$ is suppressed to minimum median values around 0.7, matching our data-informed observed posteriors.

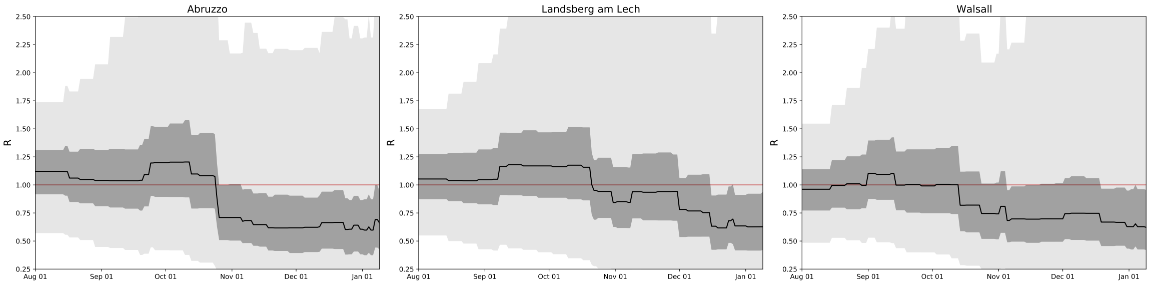

**Figure S24: Prior predictive distributions for $R_t$ of three randomly chosen regions. Solid lines indicate prior medians while shaded areas represent 95% credible intervals.**

#### 1.4. Posterior predictive distributions

The posterior predictive distribution (Figure S25) shows the predicted number of cases and deaths after observing the data. The fit is generally tight as the latent random walk allows the model to account for location-specific changes in transmission. However, note that the NPI effect parameters are shared across all locations. Therefore, our model has predictions that match the observed data and NPI effect estimates that apply across *all* of our regions. In other words, although the posterior fit is tight, our NPI effect parameters are not overfitted.

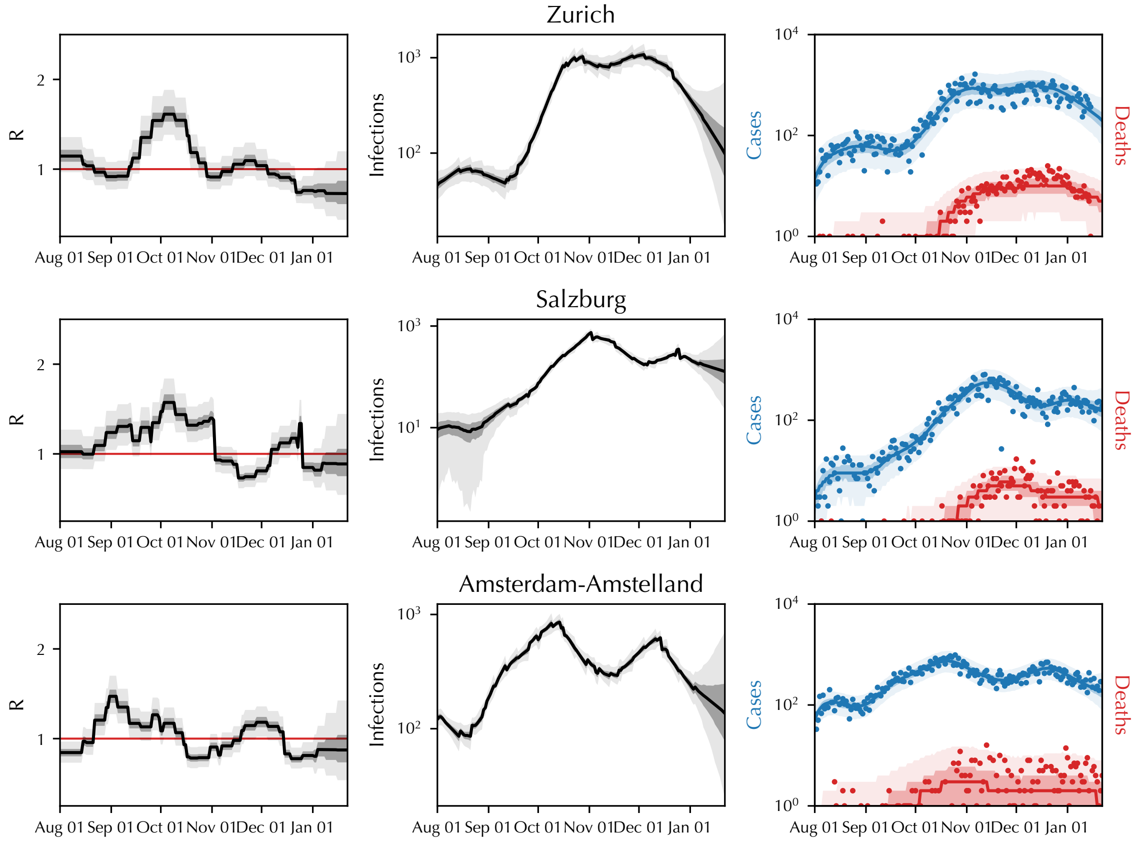

**Figure S25:** ***Left:*** **Posterior predictive distributions for three randomly chosen regions.** ***Left:*** **Inferred $R_t$.** ***Middle:*** **Inferred $N_t$ i.e., the daily number of infections.** ***Right:*** **Predicted and observed cases and deaths. Solid lines indicate posterior medians while shaded areas represent 95% credible intervals.**

#### 1.5. MCMC convergence statistics

Figure S26 shows the rank-normalized split $\hat{R}$ score (*23*) and Effective Sample Size (ESS) MCMC convergence statistics.

Values of $\hat{R}$ close to 1 indicate convergence of the MCMC sampling algorithm. Vehtari et al. suggest a sample should only be used if $\hat{R} \leq 1.01$ (*23*). Therefore, Figure S26 (left) indicates that our posterior has converged and may be used to draw inferences.

We used 250 warmup samples and 1250 iterations per chain, giving 5000 posterior samples. Inspecting Figure S26 (right), the relative effective sample size exceeds 0.5 for the vast majority of parameters, indicating low autocorrelation and quick mixing.

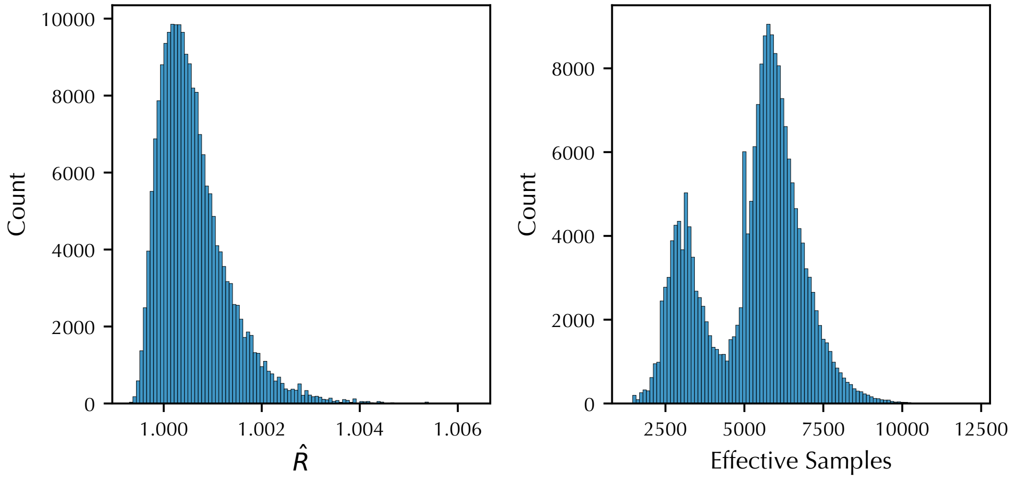

**Figure S26: MCMC convergence statistics, taken from a run using the default model with default settings and values for all parameters. *Left:* $\hat{R}$ values are close to 1, indicating convergence. *Right:* Effective sample size.**

#### 1.6. Meta analysis of individual runs

To explore issues around the identifiability of effect sizes at individual local areas versus across all areas, in Figure S27, we provide a meta-analysis of the effects estimated by running an individual model for each local area. In other words, we run our model for each location separately and aggregate the results by performing a meta-analysis. Specifically, we first estimate the effectiveness of interventions in each location. Then, we compute the median effectiveness of each intervention for each location. We then summarise the median effect sizes across the 114 individual runs by computing the median of the median effects and the 95% interval of the median effects.

We find that this approach gives NPI effects that are broadly similar to performing joint inference in all locations. Although the overall median effects for the closure of non-essential businesses and the banning of gatherings are lower in the meta-analysis, they remain the NPIs with the strongest effects, as in the main model run.

We note that the relative impact of different interventions cannot be disentangled for a single local area treated in isolation. However, when looking across multiple countries, an identifiable and robust effect of the interventions can be estimated.

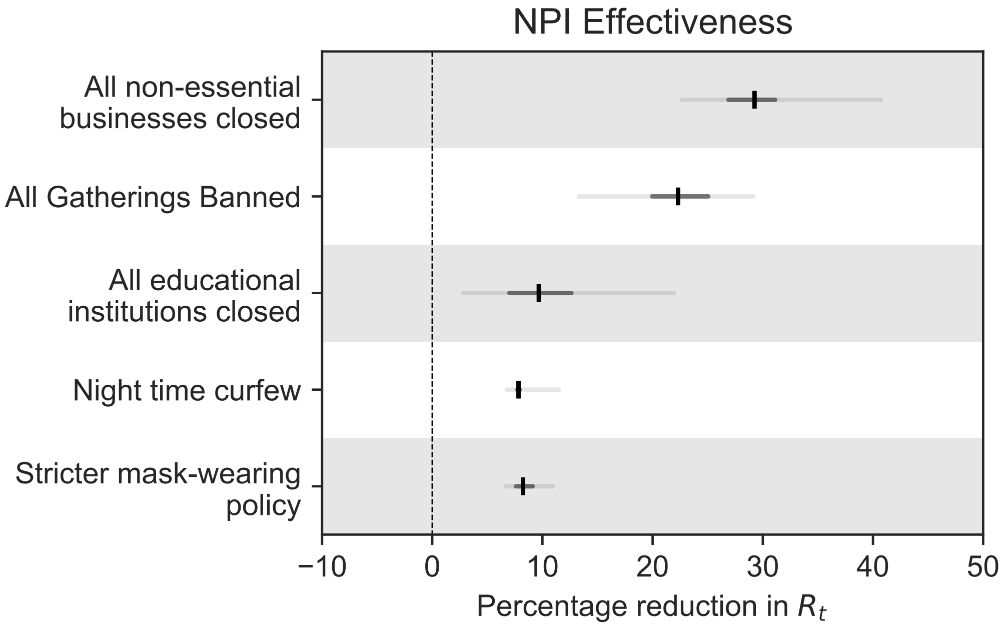

**Figure S27: Inferred intervention effect sizesn using a meta-analysis across 114 individual models; one seprate model for each local area.**

### 2. Additional results

#### 2.1. Co-occurrence of NPIs

Figure S28 shows the total number of days across all regions available to distinguish NPI effects. For every pair of NPIs (row - column), the entry shows the number of days on which only one of the NPIs was implemented (but not both or neither). Note that we do not show the traditional collinearity statistics (variance inflation factors and data correlations) since their applicability to time series data is limited. In particular, the value of these statistics in our data increases as data for a longer period becomes available, which would misleadingly suggest that we could address problems from collinearity by using less data.

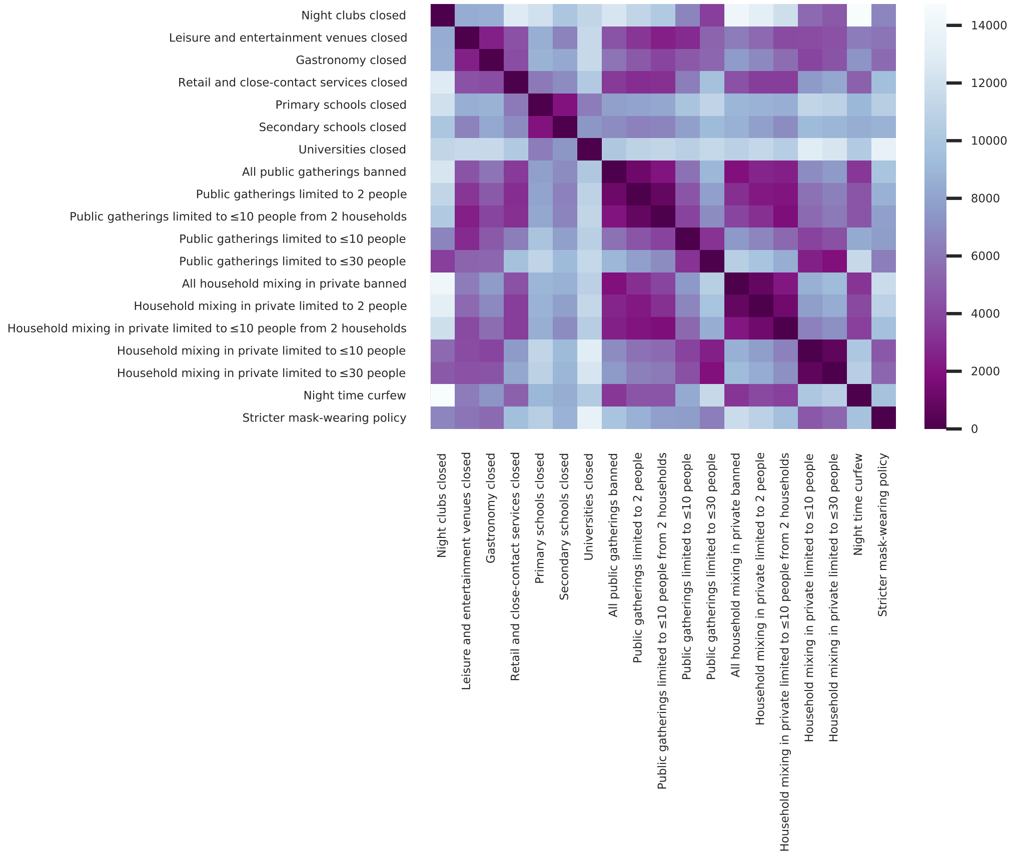

**Figure S28: Number of days across all regions available to distinguish NPI effects. For every pair of NPIs (row - column), the entry shows the number of days on which exactly one of the two NPIs was active.**

#### 2.2. Correlations between effectiveness estimates

The mean effect parameters $\beta_i$ of NPIs that are often used together are typically negatively correlated with each other, reflecting uncertainty about which NPI reduced $R$. Excessive collinearity in the data would result in wide posterior credible intervals with strong correlations (*16*). We, however, generally find weak posterior correlations between effectiveness estimates with some exceptions–see Figure S29. These weak correlations are one indicator that collinearity is manageable with our dataset. Note that we present the correlations between *marginal* effect parameters. For example, the feature household mixing in private limited to ≤10 people is made up of two marginal features: household mixing in private limited to ≤30 people and the additional effect of limiting household mixing in private to ≤10 people.

We find strong negative correlations between: Public gatherings limited to 2 people and Public gatherings limited to 2 households (-0.44); All household mixing in private banned and Household mixing in private limited to 2 people (-0.45); Household mixing in private limited to 2 households and Household mixing in private limited to 2 people (-0.54), and Household mixing in private limited to ≤10 people and Household mixing in private limited to ≤30 people (-0.42). All other correlations between pairs of NPIs have an absolute value less than 0.4.

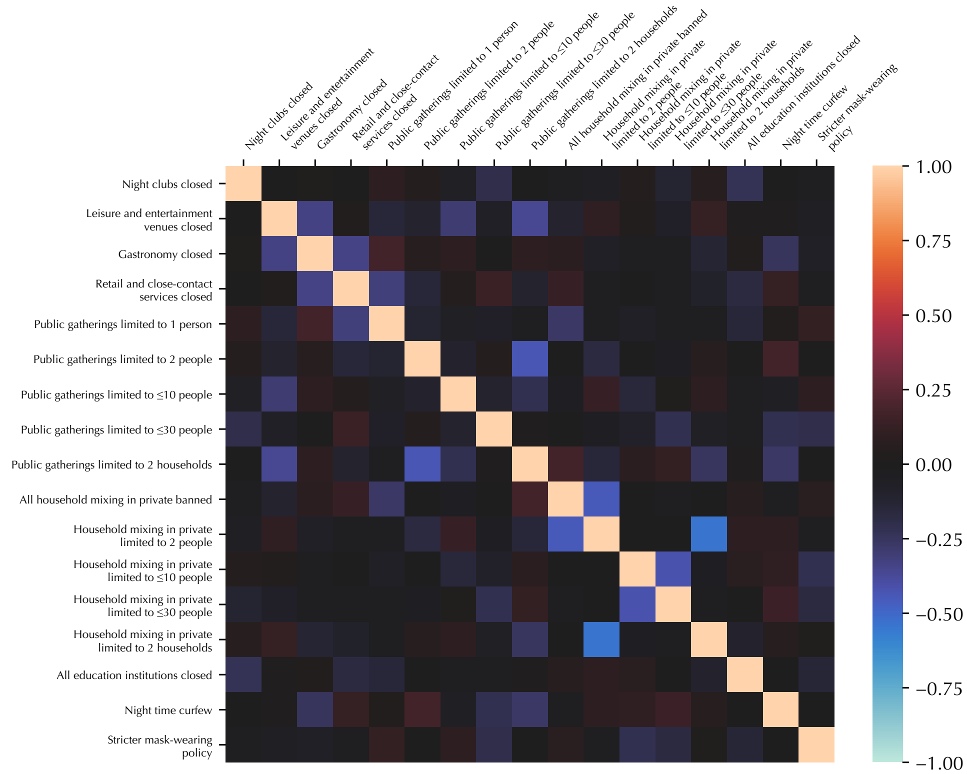

**Figure S29: Posterior correlations between effect parameters $\beta_i$.**

#### 2.3. Random walk noise scale posterior

Figure S30 shows the posterior (using a kernel density estimate) of the random walk noise scale, $\sigma_R$. We observe that the posterior is peaked despite the wide prior, suggesting that our data is informative about this parameter.

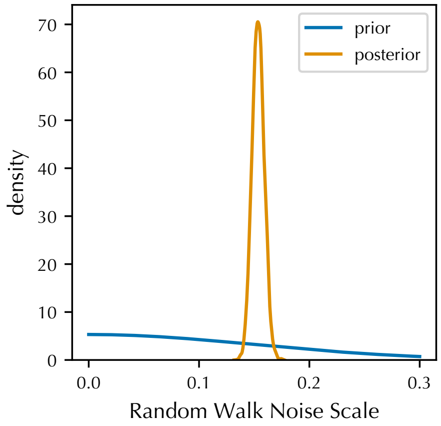

**Figure S30: Prior and posterior distributions over the random walk noise scale parameter, $\sigma_R$.**

### 3. Data details

#### 3.1. Feature creation

In this section, we describe the preprocessing steps used to create the NPI features used in our model from the raw data. All NPIs not mentioned below are extracted from the raw data in a straight-forward manner.

*Universities closed.* Some of the regions of analysis did not contain universities. For these, the “universities closed” feature was 1 (“on”) throughout the period of analysis.

*Restrictions on gatherings and household mixing.* The dataset contains the exact number of people and households allowed at various gathering types over time. To create binary features from the limitations on gatherings, we thresholded gathering limits into “gatherings limited to ≤30 people”, “gatherings limited to ≤10 people”, “gatherings limited to 2 people or fewer”, “all gatherings banned”. These thresholds were chosen based on the histogram of gathering limits to reduce collinearity between the thresholded features. Concretely, we chose these thresholds as each of the following were active for a similar number of days across all regions:

- "gatherings with >30 people are banned, but gatherings with >10 people are allowed"
- "gatherings with >10 people are banned, but gatherings with >2 people are allowed"
- "gatherings with >2 people are banned, but gatherings with 2 people are allowed"
- "all gatherings are banned"

The dataset records restrictions for 4 types of gatherings separately: public indoor gatherings, public outdoor gatherings, household mixing in private indoors, and household mixing in private outdoors. For both public gatherings and household mixing in private, there was very high collinearity between rules for indoors and outdoors. We thus aggregated the indoor and outdoor data into one NPI feature used for modelling. In the rare cases in which there were differences between rules for indoor and outdoor settings, we counted the rules for indoor settings, as indoor gatherings are expected to carry a higher risk of virus transmission.

For each entry, the dataset contains not only limits on the number of people at gatherings, but also any limits on the number of different households that these people were allowed

**Table S2: Stringency levels of mandatory mask-policies used in the dataset. The stringency level definitions were taken from the Oxford COVID Government Response Tracker (OXCGRT) dataset (*24*) (OXCGRT codebook here).**

| Stringency level | Definition |
|---|---|
| 0 | No policy. |
| 1 | Recommended. |
| 2 | Required in some specified shared/public spaces outside the home with other people present, or some situations when social distancing is not possible. |
| 3 | Required in all shared/public spaces outside the home with other people present or all situations when social distancing is not possible. |
| 4 | Required outside the home at all times regardless of location or presence of other people. |

to be from. The only such limit that was common across several countries was a limit of 2 households for gatherings of 6 people, and therefore this is the only household limit we include as a feature and whose effect we estimate.

*Mask-wearing policy.* During the data collection, we categorised mask-wearing policies into five stringency levels (Supplementary Table S2). All regions of analysis in our dataset had policies with at least a stringency level of 2 at all times during our analysis period. However, many regions switched from a level 2 policy to a policy of stringency level 3 (or higher) during our analysis period. We thus defined our mask-wearing policy feature to be 1 ("on") if the stringency level was 3 or higher, and 0 ("off") otherwise. Our inferred effect thus corresponds to the added benefit of a policy of level 3 (or higher) over a level 2 policy.

*Business closures.* We collected data on the closures of four major types of face-to-face businesses (night clubs and related, leisure time and entertainment venues, gastronomy, retail and close-contact services). To verify that we had not missed any major business type, we also collected an NPI called "all non-essential businesses closed". The data collectors were not informed that this NPI would be used for this validation purpose. The NPIs had the following definition: *"A country has suspended the operations of many face-to-face businesses. By default, face-to-face businesses are suspended unless they are designated as essential (whitelist). The whitelist is determined by each country and is different from country to country."* After the data collection, we evaluated the consistency between the business NPIs. Indeed, "all non-essential businesses" (as per the dedicated NPI) are closed whenever the four specified business types are closed, and vice versa.

#### 3.2. Data tables

**Table S3: The regions of analysis in each country and sources for daily case and death data. All links are clickable.**

| Country | Regions of analysis | Number of regions of analysis | Sources for case and death data |
|---|---|---|---|
| Austria | States | 9 (whole country) | https://www.data.gv.at/katalog/dataset/covid-19-zeitliche-darstellung-von-daten-zu-covid19-fallen-je-bezirk/resource/9eb08d45-ff99-40f1-90cd-7b3659b0bc8d |
| The Czech Republic | Administrative regions | 14 (whole country) | https://onemocneni-aktualne.mzcr.cz/api/v2/covid-19 ; Spreadsheet name: COVID-19: Přehled epidemiologické situace dle hlášení krajských hygienických stanic podle okresu (engl: COVID-19: Overview of the epidemiological situation according to the reports of regional hygienic stations by district) |
| England | NUTS 3 statistical regions | 15 (stratified random sample) | https://coronavirus.data.gov.uk/ ; Lookup table for conversion from LAU-1 to NUTS3 here. |
| Germany | Districts [including "rural districts" (Landkreise) and "urban districts" (kreisfreie Städte/Stadtkreise)] | 15 (stratified random sample) | https://github.com/jgehrcke/covid-19-germany-gae |
| Italy | Administrative regions (With the exception of the Trentino-Alto Adige region, which was split into the autonomous provinces Trentino and Alto Adige). | 21 (whole country) | https://github.com/pcm-dpc/COVID-19/blob/master/dati-regioni/dpc-covid19-ita-regioni.csv |
| The Netherlands | Safety regions | 25 (whole country) | https://coronadashboard.government.nl/veiligheidsregio/VR13/positief-geteste-mensen |
| Switzerland | Cantons | 15 (stratified random sample) | https://github.com/openZH/covid_19 |

**Table S4: Regions of analysis from each country. Local language names are given in parentheses if they differ from the English names. Abbreviations: CC: county council, RD: rural district, UD: urban district**

| Country | Regions of analysis |
|---|---|
| Austria | Burgenland, Carinthia (Kärnten), Lower Austria (Niederösterreich), Salzburg, Styria (Steiermark), Tyrol (Tirol), Upper Austria (Oberösterreich), Vienna (Wien), Vorarlberg |
| The Czech Republic | Central Bohemian (Středočeský), Hradec Králové (Královéhradecký), Karlovy Vary (Karlovarský), Liberec (Liberecký), Moravian-Silesian (Moravskoslezský), Olomouc (Olomoucký), Pardubice (Pardubický), Plzeň (Plzeňský), Prague (Hlavní město Praha), South Bohemian (Jihočeský), South Moravian (Jihomoravský), Ústí nad Labem (Ústecký), Vysočina (Vysočina), Zlín (Zlínský) |
| England | Brighton and Hove, Buckinghamshire CC, Coventry, East Derbyshire, Enfield, Essex Haven Gateway, Gloucestershire, Greater Manchester South West, Lincolnshire, North Yorkshire CC, Portsmouth, Redbridge and Waltham Forest, Southampton, Southend-on-Sea, Walsall |
| Germany | Aschaffenburg (RD), Breisgau-Hochschwarzwald (RD), Donau-Ries (RD), Ennepe-Ruhr-Kreis (RD), Enzkreis (RD), Fürth (UD), Gifhorn (RD), Hildesheim (RD), Landsberg am Lech (RD), Nuremberg (Nürnberg, SD), Minden-Lübbecke (RD), Mönchengladbach (SD), Münster (SD), Rems-Murr-Kreis (RD), Rhein-Kreis Neuss (RD) |
| Italy | Abruzzo, Aosta Valley (Valle d'Aosta), Apulia (Puglia), Basilicata, Calabria, Campania, Emilia-Romagna, Friuli Venezia Giulia, Lazio, Liguria, Lombardy (Lombardia), Marche, Molise, Piedmont (Piemonte), Sardinia (Sardegna), Sicily (Sicilia), South Tyrol (Alto Adige), Trentino, Tuscany (Toscana), Umbria, Veneto |
| The Netherlands | Amsterdam-Amstelland, Brabant-Noord, Brabant-Zuidoost, Drenthe, Flevoland, Friesland (Fryslân), Gelderland-Midden, Gelderland-Zuid, Gooi en Vechtstreek, Groningen, Haaglanden, Hollands Midden, IJsselland, Kennemerland, Limburg-Noord, Midden- en West-Brabant, Noord- en Oost-Gelderland, Noord-Holland-Noord, Rotterdam-Rijnmond, Twente, Utrecht, Zaanstreek-Waterland, Zeeland, Zuid-Holland-Zuid, Zuid-Limburg |
| Switzerland | Aargau, Basel-City (Basel-Stadt), Basel-Country (Basel-Landschaft), Friburg (Freiburg), Geneva (Genéve), Grisons (Grischun), Jura, Lucerne (Luzern), Neuchâtel, St. Gallen, Thurgau, Valais, Vaud, Zug, Zurich (Zürich) |

### 4. Additional explanations and judgement calls in data collection

Throughout the data collection process, our researchers consistently applied the definitions for each of the NPIs when determining how to code each location and period. In certain cases, however, the way NPIs were defined or implemented in a country left room for some ambiguity about how to correctly code a data point. In such situations, our researchers discussed the issue internally to ensure that all similar cases were treated similarly across countries. In certain cases, where the ambiguity could not be immediately resolved, we consulted professional or personal contacts from the respective country.

This appendix outlines some common questions and edge cases that arose during the data collection process, including detailed descriptions about how all such cases were handled.

#### 4.1. Non-mandatory recommendations

As it is very hard to consistently define various kinds of advice and non-mandatory recommendations, we only record mandatory interventions, not recommendations. Given the intractability of determining the extent to which mandatory NPIs were enforced, we did not actively research the level of enforcement. However, some regions do not have the legislative capacity to regulate gatherings in private homes. As an exception, we therefore also recorded prohibitions against private gatherings that were not legally binding. This was applicable in Italy, Austria, and the German state Northrhine-Westphalia. In each case, we confirmed with locals that these restrictions were followed by the vast majority of the population.

##### *4.1.1. Exceptions*

In Italy, due to the government's inability to introduce a mandatory limit on gatherings in private homes, a strong recommendation for a limit of 6 non-cohabiting individuals gatherings in private homes was instead issued on 13th October 2020. Private parties in bars, restaurants and other public places were also prohibited, and weddings and other ceremonies had guest limits reduced to 30. According to Prime Minister Conte, *"We can't introduce a binding rule, but the message we must send at all costs is this: do not organise private events with lots of people."* It seems they could not legislate for this rule of 6 in private homes but the strong messaging and quote seems that it would have altered public behaviour to try and stick to this rule, at least in the most part. Also: *"The wording of the decree suggests that you can't invite more than six guests, not that there can't be more than six* *people in total."* (https://www.thelocal.it/20201019/covid-19-what-to-know-about-italys-new-rule-of-six) However, *"It's not unambiguous, though, and for simplicity's sake the advice has been widely described in the Italian media as 'no more than six to dinner."* Keeping this in mind, we judged that it was best to code the limit on people as 6, and this judgment was validated in conversation with an Italian epidemiologist.

#### 4.2. Exceptions to rules on gatherings

Sometimes gathering restrictions don't apply to certain types of gatherings (e.g. not to funerals) or types of areas (e.g., parks in the UK), or apply only to certain areas (e.g. parks in Switzerland). Since our NPIs are defined in terms of "most or all" of the population/venues/events being affected, we used our best judgment to determine whether this was the case. E.g., when only funerals are exempt from a gathering ban, then certainly the limit applies to most gatherings, as funerals are quite rare. However, when all "social and cultural events" are exempt, the limit cannot be said to apply to "most gatherings", and we would not record it.

##### *4.2.1. Examples*

In Austria, the state Burgenland seems to have had stricter measures than the national measures between 2020/10/23 and 2020/11/02 (national measures were significantly tightened on 2020/11/03). However, these seem to have only applied to sporting events. (See the State ordinance, archived corona-ampel.gv.at, the press release on the State authority's website.) Since the measures only applied to sporting events, not to all or most events, we did not record a state-level deviation in any NPIs related to this.

In England, from the 3rd of July onwards, gatherings were only allowed for up to thirty people. There were quite a few exceptions for public gatherings. Exceptions (note: all three of the bullet points below must be satisfied):

- (i)the gathering has been organised by a business, a charitable, benevolent or philanthropic institution, a public body, or a political body,
- (ii)the person responsible for organising the gathering ("the gathering organiser") has carried out a risk assessment which would satisfy the requirements of regulation 3 of the Management of Health and Safety at Work Regulations 1999(1), whether or not the gathering organiser is subject to those Regulations, **and**

- (iii)the gathering organiser has taken all reasonable measures to limit the risk of transmission of the coronavirus, taking into account the risk assessment carried out under paragraph (ii).

These exceptions would have to be approved by the government and were granted larger gatherings with the numbers decided on a case-by-case basis. As most public gatherings did not warrant applying for an exception, we recorded this as a maximum of 30 for both public and private outdoor spaces.

In Enzkreis, Baden-Württemberg, Germany: on 24/10/2020 gatherings and private events were limited to 5 individuals except in cases of meetings with family members or just one other household. This was coded as a public/private as well as indoor/outdoor restriction on gatherings of 5 individuals, with no household limit.

In Germany, North Rhine Westphalia had a long list of restrictions for meetings. Meeting with $\oint$10 people was banned, but events with safety restrictions (including either fixed seating or 1.5m distance) could get around this. There was a requirement to apply for permission for events with $\oint$300, to verify that hygiene protocol was being followed. The rule stated:

- **§ 1, paragraph 2: Several people may only meet in public space if they are**<br>1. exclusively to relatives in a straight line, siblings, spouses, life partners, 2. exclusively about people from a maximum of two different domestic communities, 3. to accompany minors and persons in need of support, 4. for absolutely necessary meetings for reasons relevant to care or 5. in all other cases by **a group of no more than ten people**. ... Other gatherings and get-togethers of people in public spaces are not permitted until further notice; **with exception of**:
- 1. unavoidable accumulations during the intended use of permitted facilities (in particular when using passenger transport services and its facilities),
- 2. **Participation in events and meetings permitted under this Ordinance**,
- 3. Permissible sporting activities and permissible youth work and youth social work,
- 4. Compulsory meetings for professional practice in public spaces. ...
- **At events and meetings** that do not fall under the special provisions of this ordinance, **suitable precautions must be taken to ensure hygiene, to control access, to ensure a minimum distance of 1.5 meters (also in queues) between people who do not belong to the requirements specified in § 1, paragraph 2, and, if necessary, to implement an obligation to wear a face-to-face mask** (Section 2, paragraph 3). Apart from outdoors, easy traceability must also be ensured in accordance with Section 2a (1).

**If the participants sit in fixed places during the event or meeting, the requirement of a minimum distance of 1.5 meters between people can be replaced by ensuring the special traceability** according to § 2a paragraph 2.

- (2) **Events and assemblies that do not fall under the special provisions of this ordinance with more than 300 participants require a special hygiene and infection protection concept** in accordance with Section 2b, which at least ensures the requirements of Paragraph 1. ...
- (4) Notwithstanding paragraphs 1 and 2, **large festive events are prohibited** ...
- (5) Paragraphs 1 and 2 do not apply to **parties** (events with a primarily social character). These **are only permitted for an outstanding occasion (e.g. anniversary, wedding, christening, birthday, graduation party) and with a maximum of 150 participants. The distance requirement and an obligation to wear a face-to-face mask do not apply** as long as suitable precautions for hygiene and easy traceability are ensured

We classified this as a 10 person limit on public gatherings and a 150 person limit on private gatherings (as nearly all private events with 150 people will be due to an "outstanding occasion" and so nearly no private events with ∭150 people were actually banned). As the 150 person limit was reduced, we reduced the limit on private gatherings accordingly. When similar situations came up in other regions, we followed a similar solution.

In the Czech Republic, a national measure says basically that ~all events and gatherings are limited to 1000 people outdoors and 500 indoors, except for ~events in "structurally partitioned facilities." The full quote is *"this prohibition does not apply to... large events held in structurally partitioned facilities (sports stadiums, exhibition grounds, etc.) with the presence of more than 1,000 people at any one time if the event is held largely outdoors, or more than 500 people in each of a structurally separate section (by means of a mobile barrier, e.g. portable fence) of a facility if the event is largely held indoors, provided the entire facility is divided into a maximum of five structurally partitioned sections, each having access from outdoors and the attendees being unable to move between the sections; the distance between people in adjacent sections must be at least four metres; in premises with seating (e.g. sports stadium stands, etc.), attendees of the large event must be seated in only every second row and must be separated from other attendees in these rows by at least one empty seat"* and, also excluded are *"meetings, conventions and similar events held by constitutional bodies, public authorities, courts and other public entities, which are held by law,"* and *"gatherings pursuant to Act No. 84/1990 Coll., as amended, on the Right to Gather."* While this was a borderline case, we concluded that "most" events above that size were prohibited (and the bigger events that were allowed to occur with barriers could be considered multiple events of only 1000/500 (outdoor/indoor) people), and recorded maximum gathering sizes of 1000/500/1000/500 for public outdoor/public indoor/private outdoor/private indoor gatherings, respectively.

In Switzerland, until October, there was a limit on the total number of attendees at an event as well as a limit on divisions ("Sektoren") within events where it might not be possible to social distance for the entire event. We used these division limits as gathering restrictions in the absence of other more specific restrictions.

###### 4.2.2. *Exceptions*

In Austria, the Government advice page states that private Christmas parties of 10 people from 10 different households were allowed on December 24 and 25. The regulation attached to this advice allows meetings of 10 people from 10 different households. Therefore, this was taken to mean that public outdoor/indoor and private outdoor events were limited to 10 people on the 24th and 25th of December.

In Germany, over Christmas (24/12-26/12), one was allowed to meet up to four family members regardless of the number of households. This was coded as a limit of 5 with no household restriction (5,no).

##### 4.3. Special arrangements for educational institutions

In some cases, special teaching arrangements (e.g., online teaching or an option to keep children home from school) made it ambiguous whether schools should be recorded as open or closed. As a general rule, we aimed to determine whether it was the case that "most or all" (i.e., ∮50%) of children at a given educational level (i.e., primary, secondary, or university) were in school. Below is a list of examples from various countries and explanations of our decision criteria.

###### 4.3.1. *Examples*

In Lower Saxony, Germany, parents were allowed to keep their children home from the 14th to the 18th (the last week of school before holidays). This was coded as no closure on schools.

In North Rhine Westphalia, Germany, during the last week before christmas, parents of school children in grades 1-7 (ages 7-13) could choose to keep their children home, while grades 8-13 (ages 14-19) had to do online schooling. We coded this as a restriction on secondary schools but not on primary schools, after consulting an online document which records what fraction of pupils actually attended school `https://www.schulministerium.nrw.de/system/files/media/document/file/Zeitreihe-Kreise-KW51.pdf`.

In Minden-Lubbecke, Germany, the largest University started its normal face-to-face courses on the 2nd November. However, they hint that there might have been mandatory face-to-face introductory events earlier. In addition, a few of the courses involved working for part of the year at companies and followed a different schedule. (See here: `https://www.fh-bielefeld.de/minden/informationen-fuer-studierende-und-erstsemester`). Consequently, the end of autumn vacation was coded as 1st November.

In Furth, Germany, a small university's "exam time" extended into August (see here). However, this was counted as part of the summer holidays, as it is not clear how many students had exams in August.

In Portsmouth, UK, the University of Portsmouth had a "Consolidation Week" during term. While classes were not in session, students most likely remained on campus and continued using the facilities (to the extent that they had been doing this before). Consequently, we did not consider this as a closure of the university.

In Portsmouth, UK, the University of Portsmouth required international students to commence one week before the rest of the students. However, as it seems like internationals make up just ~4000 of the 25,000 students, we used the date when most (i.e., including nationals) students began term.

In the Czech Republic, national rules for primary schools involved a long period where 1st and 2nd grade pupils (1st and 2nd years of primary school) were going into school, and the older grades (3rd, 4th, and 5th years of primary school) were not. As attendance was roughly 40% (2 of 5 years attending), we did not record an NPI for such periods.

Similarly, in the Czech Republic, national rules for secondary schools involved periods where grades 10-13 (the last 4 grades of secondary school) were closed, and grades 6-9 (the first 4 grades) went into school on alternate weeks (i.e., if they followed the rules exactly, you would expect 50% attendance). Since about 4/8 * 0.5 = 25% of secondary school students were attending school at any one time, we counted an NPI for periods where grades 10-13 were closed and grades 6-9 attended on alternate weeks, as most students were likely not in attendance.

In Austria, the school situation was somewhat complicated. There were plenty of provisions for students to go into school after the schools had been “closed” or switched to “distance learning”. The key periods are: Lockdown Light 1 (2020/11/03-2020/11/16), Lockdown Light 2 (2020/12/07-2020/12/23), Strict Lockdown 1 (2020/11/17-2020/12/06), and Strict Lockdown 2 (2020/12/26-post our data window). During the ‘light’ lockdowns, only ~"higher schools" were distance learning, which roughly corresponds to Sekundarstufe II on this page. During the strict lockdowns, all schools were distance learning. We found two relevant sets of data regarding school attendance: a comprehensive survey on 2020/11/17 as reported here, and another survey on 2020/11/27 reported here. The 2020/11/17 survey claims an average attendance of 15% across all school years nationwide, ~23% (inferred) average attendance in primary schools nationwide, and 25% attendance across all school years for regions with the highest attendance. The 2020/11/27 survey has similar numbers. It claims that the all-age nationwide average attendance rate increased by 2%, from 15% to 17%. Based on this, we infer that in Strict Lockdown 1 our secondary school NPI was in effect in all regions, while our primary school NPI was more borderline: Oberoesterreich and Niederoesterreich had an overall attendance of 25%, and the numbers from the other regions suggest that this would have been concentrated in primary schools. We estimated that there was ~32% attendance in primary schools in Oberoesterreich and Niederoesterreich. Based on this, we decided to code the primary school NPI as active, since assuming any specific regional variation would be based on speculation and because we expect the regional variation to be small (all regions had attendance fairly close to 30%). For Strict Lockdown 2, we did the same as for Strict Lockdown 1. We did this because the rules were similar and because this lockdown was only relevant for a period of 3 days, since schools were on holiday until 2021/01/07. For the light lockdowns, it was clear that we should not record an NPI for primary schools, as they operated as normal. We did not record an NPI for secondary schools either, as Sekundarstufe II was distance learning and the rest of secondary school-age students, Sekundarstufe I, were going to school as normal (Sekundarstufe II covers 5 grades, Sekundarstufe I covers 4 grades. If we assume that all of the Sekundarstufe I students went into school, all of the Sekundarstufe II students stayed at home, and that all year groups had the same number of students, then 4/9 = 44% of students attended school). In summary, we coded both school NPIs as active for the strict lockdowns and inactive for the light lockdowns and any other non-holiday periods.

##### 4.4. Distinctions between types of gatherings (indoor/outdoor, private/public, number of households)

In some cases, countries did not clearly distinguish between indoor/outdoor and private/public gatherings. In others, their rules did not map neatly onto our distinctions. We evaluated such situations on a case-by-case basis while striving to ensure consistent handling across all countries and cases.

###### *4.4.1. Examples*

In England, public indoor gatherings are not defined distinctly from private indoor gatherings in the legislature. The UK seems to use "indoors" for all categories but has clear exceptions where things are allowed (prison, education, etc). While some public buildings were open in the second lockdown, it does not appear that people were able/supposed to organise gatherings in them. This was vague in the sense that there was leeway for students to meet and study in libraries together in groups, and it was not illegal for families to go to shopping malls together. We concluded that legislation on "indoor gatherings" applies to both public and private. *Ref:* (pages 9-16) `https://www.legislation.gov.uk/uksi/2020/1611/pdfs/uksi_20201611_en.pdf`.

Regarding household limits in England prior to the introduction of the tier system on the 14th of October: this (as well as a key amendment that introduced the rule of six) was the key legislation that affected gatherings. Neither legislation mentions anything about a maximum number of households. However, some sources had vague messages about government guidance (e.g., a government news release that explained the rule of six stated this: *The rule applies across England and replaces the existing ban on participating in gatherings of more than 30 and the current guidance on allowing two households to meet indoors."*). Upon further investigation, it appears that government guidance and legislation differed:

- (published on September 14) *"Since July, the Government guidance has stated that no more than six people should meet outside, while only two households are allowed to meet indoors. However, this was different from the actual law, which stipulated that any gathering over 30 people was illegal and could be broken up by police."*
- (A BBC article referencing the same thing, published September 9) *"At present, the guidance says two households of any size are allowed to meet indoors or outdoors, or up to six people from different households outdoors. Until now the police have had no powers to*

*stop gatherings unless they exceeded 30 [referring to the amendment which instated the rule of six]."*

From this, it seems that the Government was encouraging a limit on households and meetings indoors/outdoors, but this limit was not mandatory. The guidance would dictate that we code "any number of people from up to two households (no,2) for all indoor and outdoor gatherings", and "six people from any number of households (6,no) for outdoor gatherings", but the legislation dictates that we only code (30,no). As the guidance was not mandatory, we coded in accordance with the legislation: (30,no).

In North Rhine Westphalia, Germany, there were never any laws about meeting inside homes. However, there was a strong nationwide policy that all gatherings, including those inside homes, should be limited to a maximum of 5 people and a maximum of 2 households. Consequently, this was coded as a (5,2) restriction on private outdoor and indoor spaces.

In Italy, a private indoor and outdoor gathering restriction was introduced on December 24 where 2 people were allowed to travel to another home to meet other people. Quote: *'It is possible to visit one other private residence within the same region between 5am and 10pm, no more than once a day. Curfew must be respected. A maximum of two persons may make such journeys, plus any children in their household under the age of 14.'* Assuming that the average household has 2 people, we recorded the limit at 4.

In Switzerland, in the absence of a private gathering limit, there was a private event limit of 100 for events with masks aside from when seated and eating, and a limit of 15 for events without masks. We coded this as a limit of 100 people, as we considered mask-wearing at such private events (especially where eating and drinking happens) unlikely to be strictly enforced.

In Bavaria, Germany, between August and October, one was allowed to meet up to 10 individuals or individuals from only one further household in public places. This was coded as up to 10 with no household restriction (10,no).

Regarding private gathering sizes during stay at home orders in the Czech Republic: there were periods where people were ordered to stay at home except for essential journeys. There were meeting limits for public places (at some times the limit was 2, and at others it was 6). There were limits for private places. On the face of it, you might think that meeting in private was prohibited except for essential reasons. However, according to a Czech national we spoke to, people took the private gathering limit to be the same as the public counterpart. And public statements from ministers were also quite confusing (e.g. in this article a spokesperson for the Ministry of Health seems to say that private meetings have no size limit). We recorded the private gathering limit to be the same as the public one in these cases.

#### 4.5. Other edge cases

There was a range of special cases that did not neatly fall into any of the above categories. As before, we evaluated each of these situations on a case-by-case basis, while striving to ensure consistent handling across all countries and cases.

##### *4.5.1. Examples*

In Switzerland, cantonal authorities have autonomy to make decisions that overrule federal law. However, during the pandemic, they largely followed federal law, thus we deferred to federal law in cases where cantonal law did not state otherwise. Note that the current federal laws state: *"The following overview shows the rules and bans that currently apply nationwide. In other words, these measures, at the very minimum, apply throughout Switzerland. Stricter measures may apply in certain cantons. You can find information about these in the section entitled Cantonal measures"*. Consequently, we only deferred to cantonal law where it was (a) stricter than federal law (b) explicitly stated in press releases that it would not be following federal law.

In Italy, while traffic light zones were mostly implemented by region, there was an unusual situation in Trentino-Alto Adige/Südtirol. This is because *'The region is divided into two autonomous provinces: Trentino (Autonomous Province of Trento) and South Tyrol (Autonomous Province of Bolzano)'*. Both of these have fairly similar population sizes of ~0.5m. There were cases where Bolzano was in the red or orange zone while Trento was in the yellow zone, resulting in many more NPIs being active in Bolzano compared to Trento in the same period. We contacted an Italian epidemiologist who agreed with this assessment, but who noted that *"the only exception is Trentino and Alto-Adige which are one region (Trentino Alto-Adige), but in the data are often reported as separate provinces. It is best to keep these two separate because measures are applied at the provincial level here."* Consequently, we coded these areas by the two provinces. In Baden-Wurttemberg, stay-at-home-orders applied from the 12th December. When they applied, public gatherings were generally forbidden, with the main exception being that you were allowed to exercise in the fresh air with your own household or with a single person from another household (this is very clear in the orders that applied from the 16th). We coded this as allowing public outdoor gatherings of 2 people, and public indoor gatherings of only 1.

##### 4.6. Sub-area NPI implementation

Sometimes, decisions about NPIs might be made on a level even more fine-grained than the local areas for which we collect data. For example, if a local area contains two towns, it might be the case that only one of is a hotspot and ends up introducing certain NPIs (although the other may follow suit later). In these situations, we only recorded the NPI as active if and when they affected the whole local area.

###### *4.6.1. Examples*

For example, in the area of Essex Haven Gateway, the local administrative unit of Braintree changed tiers on different dates than the two other administrative units in that area. In this case, we only coded changes in NPIs when all of Essex Haven Gateway had changed tiers.

##### 4.7. Mandatory safety measures

In some cases, it became required to implement certain safety measures for events and gatherings to occur. E.g., in Germany, cinemas operated under safety restrictions such as only allowing 25% capacity; in Italy, it was prohibited to loiter outside bars. In such cases, we did not record the safety measures and only considered actual closures and prohibitions, as nearly everything had some kind of safety measure.

###### *4.7.1. Examples*

In Austria, national rules for the maximum number of people at public events over the summer of 2020 allowed events to apply for permits to increase their capacity. For example, the default maximum number of people as of 2020/08/01 for public outdoor events with fixed seating was 750, and a permit could be granted to increase this to 1250. (See the national rules saved on web.archive.org on 2020/08/02.) Since "most or all" events were not constrained by the without-special-permission numbers (our impression is that a permit was not especially difficult to obtain), we coded the higher numbers that were allowed with special permission.

Similarly, in Austria, national rules for the maximum number of people allowed at public events over the summer of 2020 depended on whether the events used fixed seating. (E.g. see the national rules saved on web.archive.org on 2020/08/02.) Again, we used the higher, fixed seating numbers because it is not the case that "most or all" events bigger than the non-fixed-seating limit were constrained by the non-fixed-seating limit (i.e. they probably mostly had fixed seating rather than being cancelled).

In Lower Saxony, Germany, there was a rule that you could meet 10 people without any safety precautions, and as long as you keep a 1.5m distance, private gatherings and celebrations could consist of: 25 people in your home, 50 people in your garden, 100 people in any publicly accessible location *"including in rooms provided outside of one's own home and in catering establishments."* Additionally, for purely seated non-private events (e.g. cinemas), and for at least temporarily standing events with a safety concept (e.g. congresses), you could have up to 500 visitors. (Some of these numbers were later lowered due to high incidence.) Since we expect people to comply with the safety precautions in public spaces, public outdoor and indoor were coded as 10 people. Private spaces were coded as 100, since rented venues are counted as private.

#### 4.8. The timing of implementation

We record the date when an NPI is expected to have affected behaviour. If an NPI is announced to be in place immediately, the starting day is the day of the announcement, unless it was announced in the evening and clearly only in effect on the next day. One exception to this rule is nightclubs, which we counted as the day of (as most nightclub activity happens at night).

For schools, we considered the fact that such institutions are closed on weekends. Consequently, if it is announced on Thursday that schools will stay closed starting from Monday, the actual date when schools were closed is Saturday, and that is the date we recorded.

##### *4.8.1. Examples*

In Switzerland, since some laws came into place at 5pm on a certain day, we counted the next day as the first day of restriction in all of these cases.
